# Insulin resistance reverses despite weight gain with pan-PPAR agonism, revealing its plasma molecular correlates in type 2 diabetes

**DOI:** 10.64898/2026.11.06.26367120

**Authors:** Zehui Cao, Jiaqi Li, Naixin Zhang, Haonan Chen, Desi Pan, Qiaochu Chen, Yuanbang Mai, Xintong Yao, Shanyue Sun, Yaqing Liu, Wanwan Hou, Ying Yu, Yanpeng An, Ziyu Yuan, Jingze Tan, Xingdong Chen, Yan Zheng, Hao Wu, Jiucun Wang, Jun Chang, Chen Ding, Qingxia Huang, Qi Wang, Binghua Jiang, Sijia Wang, Qianqian Peng, Wenran Li, Dejing Pan, You Wang, Tao Xu, Linong Ji, Weiping Jia, Li Jin, Zhiqiang Ning, Human Phenome Project Technological Consortium, Human Phenome Project Scientific Consortium, Antonio Vidal-Puig, Xianping Lu, Leming Shi, Yuanting Zheng

## Abstract

Insulin resistance (IR) and body weight are closely linked in type 2 diabetes (T2D), so their molecular correlates are hard to separate in observational data. Pan-PPAR agonism offers a test, because it improves insulin sensitivity while body weight increases. In the ChiHOPE cohort of 835 T2D patients from two randomized phase 3 trials of chiglitazar, 53% of chiglitazar-treated patients with HOMA-IR-defined IR at baseline no longer met the IR threshold after treatment, compared with 29% with placebo and 32% with sitagliptin, while BMI rose. We integrated plasma metabolomics and proteomics into a molecular estimate of HOMA-IR. It outperformed routine clinical measures, disagreed with HOMA-IR classification in 27% of patients, and identified adverse lipid, liver and inflammatory profiles within each HOMA-IR class. With chiglitazar, the molecular estimate fell despite weight gain, with lower proteoglycan 4 and higher glutamine and glycine. Circulating markers of IR can therefore improve without weight loss.

**Highlights:**

- Chiglitazar reversed HOMA-IR-defined insulin resistance in about half of patients as BMI rises
- A plasma multi-omics score estimated HOMA-IR better than routine clinical measures
- The molecular estimate enabled better stratification by HOMA-IR
- IR-linked plasma molecules improved despite weight gain, led by falling PRG4

## Introduction

Insulin resistance (IR), impaired insulin action in multiple tissues, especially adipose tissue, liver and muscle^1–6^, is one of the earliest defects in type 2 diabetes (T2D)^7,8^ and raises the risk of its cardiovascular, kidney and liver complications^9–16^. Therefore, IR reversal is a mechanistically meaningful therapeutic goal to achieve long-term glycemic control and reduce the risk of progressive complications.

Yet the treatment of T2D is increasingly built around weight loss^17–26^. This shift has been driven by the growing global burden of obesity^27^ and the substantial weight loss achieved with newer incretin-based therapies^28–30^. Although weight loss can improve insulin sensitivity^31–43^, prediabetes remission can occur without weight loss and is accompanied by improved insulin sensitivity^44^. Diabetes also occurs in lean individuals^45,46^, particularly in East Asian populations, in whom IR and T2D develop at lower BMI^47^. These observations suggest the existence of diabetes-related IR independent of obesity. However, because body weight and IR often covary both cross-sectionally and during weight-loss interventions^32,38,48^, these settings alone cannot cleanly separate their molecular signatures. As a result, we still do not know which molecular signatures of IR reflect insulin action itself and which simply track body weight.

Peroxisome proliferator-activated receptor (PPAR) agonists with PPARγ activity can directly improve insulin sensitivity despite causing weight gain^49–57^. This weight gain is not simply an adverse effect. PPARγ activation expands the lipid storage capacity of subcutaneous adipose tissue^58–60^ and diverts lipid away from the liver and muscle, so insulin action can improve while fat mass increases^61–63^. The molecular mechanisms responsible for reversing IR by PPAR agonists are different from those of weight loss^64–66^, and separate preclinical studies have characterized distinct metabolomic changes after pioglitazone and semaglutide treatment^67,68^. PPAR agonism therefore offers what weight-loss studies cannot: a setting in which insulin sensitivity and body weight move in opposite directions.

Omics studies have advanced the molecular understanding of T2D and obesity, providing mechanistic insights into disease processes^69–73^ and treatment-associated molecular changes^74–77^. Multi-omic data can dynamically capture heterogeneous metabolic health states and intervention responses beyond traditional clinical measures, including body mass index (BMI)^73,78–81^ and chronological age^71,82,83^. Therefore, longitudinal multi-omics profiling under randomized PPAR agonist perturbation provides a framework to distinguish weight-coupled molecular signals from IR-associated abnormalities, and to define the molecular signature underlying IR reversal independently of weight loss.

Here, we used pan-PPAR agonism in two randomized trials to separate the molecular signatures of IR and body weight. Our cohort included 835 patients with T2D from two phase 3 randomized clinical trials of the pan-PPAR agonist chiglitazar, with placebo and the DPP-4 inhibitor sitagliptin as comparators^84–86^. Across more than 2,000 serial fasting blood samples collected at baseline, week 12, and week 24, we generated comprehensive longitudinal multi-omics profiles spanning genomics, methylomics, circulating miRNAomics, proteomics, and metabolomics. This design asks a question that weight-centered therapy cannot answer: what does restoring insulin action look like when it is uncoupled from weight loss? We integrated circulating metabolites and proteins into molecular estimates of HOMA-IR and followed them under randomized treatment. IR reversed in about half of treated patients with baseline IR while BMI rose. The molecular estimate outperformed routine clinical measures, exposed metabolic risk missed by fasting HOMA-IR, and showed IR-linked molecules shifting towards insulin sensitivity as weight increased.

## Results

### Chiglitazar reverses insulin resistance while BMI rises

We studied the Chiglitazar perturbed Human multi-Omics ProfilE (ChiHOPE) cohort (http://fudan-pgx.org/chihope), comprising 835 patients with T2D from two randomized, double-blind, phase 3 trials^85,86^ who consented to this companion multi-omics study (**Fig. 1A**). The ChiHOPE cohort is predominantly male (*n* = 529, 63.4%), middle-aged (median [interquartile range, IQR] = 51 [44, 58] years), and mostly obese by Asian criteria^87^ (25.7 [23.8, 27.9] kg/m^2^, *n* = 515 [61.7%] for obese individuals [BMI ≥ 25 kg/m^2^]) at baseline, with insufficient glycemic control (HbA1c ≥ 58.5 mmol/mol [7.5%] and ≤ 85.8 mmol/mol [10.0%]) despite adherence to a strict diet and exercise regimen (**Figs. 1B-C**). Patients with type 1 diabetes (T1D) or those receiving glucose-lowering treatments were excluded^85,86^ (**Methods**).

**Figure 1.**
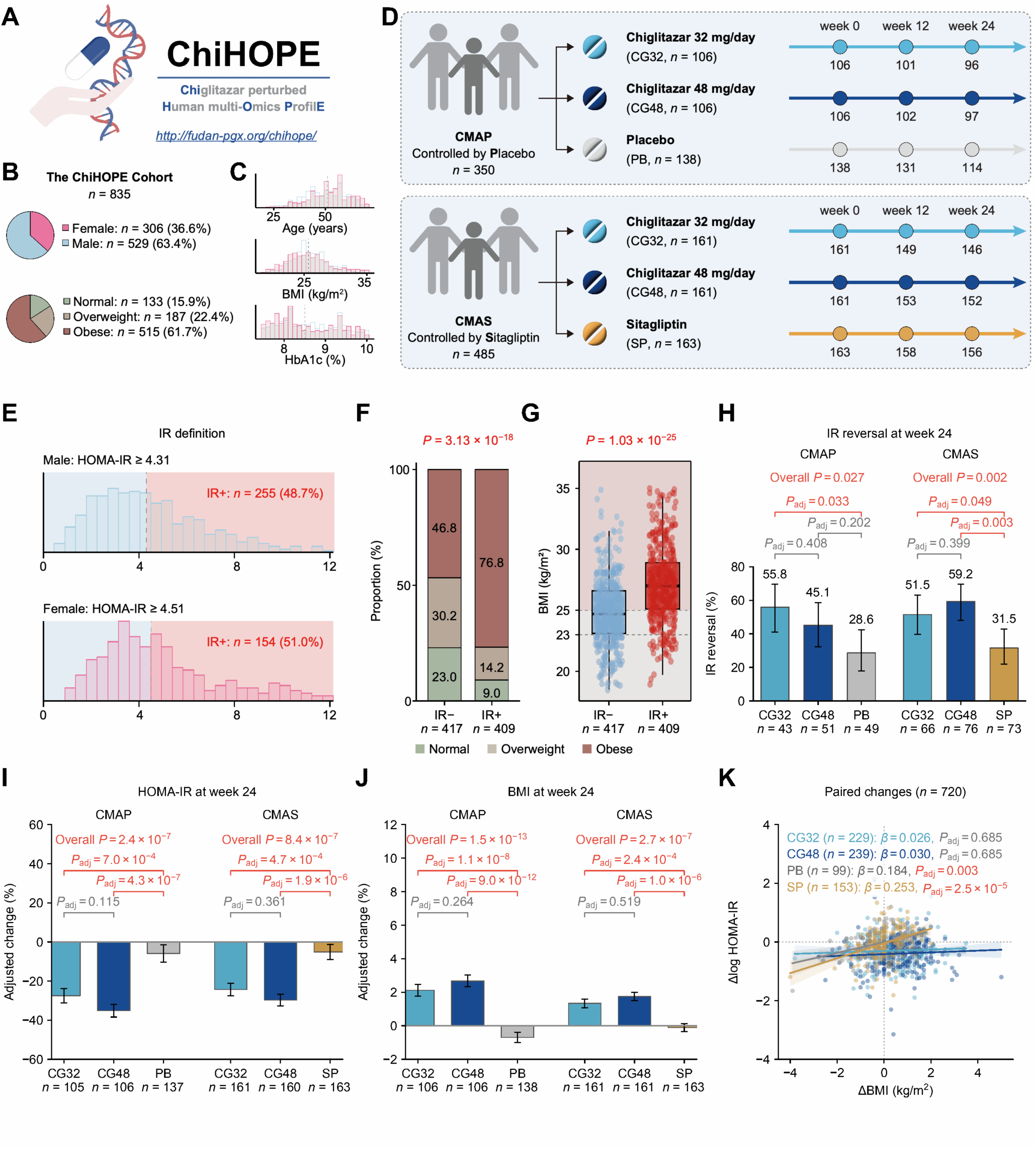
Chiglitazar reverses insulin resistance while BMI rises. (**A**) The ChiHOPE multi-omics study. (**B**) Baseline sex and BMI-category proportions in 835 ChiHOPE participants. Normal weight, overweight and obesity were defined as BMI values of 18.5–<23, 23–<25 and ≥25 kg/m^2^, respectively, according to the 2000 Asia-Pacific recommendations co-sponsored by the WHO Regional Office for the Western Pacific^87^. (**C**) Baseline distributions of age (years), BMI (kg/m^2^) and HbA1c (%) among 835 participants, colored by sex. (**D**) Treatment allocation and numbers of participants with available assessments at baseline, week 12 and week 24 in the placebo-controlled CMAP trial (*n* = 350) and sitagliptin-controlled CMAS trial (*n* = 485). (**E**) Baseline HOMA-IR distribution and sex-specific HOMA-IR thresholds used to define insulin resistance (IR): ≥ 4.31 in men and ≥ 4.51 in women^88^. (**F**) Baseline proportions of BMI categories by IR status. The overall distribution of BMI categories was compared between IR+ (*n* = 409) and IR– (*n* = 417) individuals using Fisher’s exact test. (**G**) Baseline BMI distributions by IR status. Boxes represent the interquartile range (IQR), horizontal lines indicate medians, and whiskers extend to the most extreme observations within 1.5 × IQR of the box limits. Each point represents one participant. BMI was compared between IR+ (*n* = 409) and IR– (*n* = 417) individuals using a two-sided Wilcoxon rank-sum test. (**H**) Observed proportions of participants who transitioned from IR+ at baseline to IR− at week 24 (*n* = 358). Error bars indicate 95% Wilson binomial confidence intervals. Overall differences among the three treatment groups were assessed using Fisher’s exact tests separately within CMAP and CMAS. Pairwise differences were assessed using two-sided Fisher’s exact tests, with Holm correction across the three comparisons within each trial. (**I**-**J**) Model-estimated percentage changes from baseline to week 24 in HOMA-IR (**I**) and BMI (**J**), shown separately for CMAP and CMAS. Bars and error bars represent estimates and delta-method SEs. Analyses used all eligible available week 0, week 12 and week 24 observations for each outcome separately. Separate REML linear mixed-effects models were fitted within each trial to log-transformed outcomes. Models included categorical visit, treatment arm, their interaction, sex and age, with a participant-specific random intercept. Overall differences in changes among the three treatment groups were assessed using F-tests from linear mixed-effects models separately within CMAP and CMAS. Pairwise differences were assessed using two-sided t-tests, with Holm correction across the six comparisons (three treatment-group comparisons at weeks 12 and 24) within each trial and outcome; only week-24 results are shown. (**K**) Associations between changes in BMI and log-transformed HOMA-IR from baseline to week 24 among 720 participants with paired measurements. Each point represents one participant; lines and shaded areas represent adjusted regression lines and pointwise 95% confidence intervals. Linear regression models included BMI change, treatment group, their interaction, sex, age and clinical trial. Arm-specific associations were assessed using two-sided *t*-tests, with Holm correction across the four treatment groups. *β* represents the change in log HOMA-IR per 1 kg/m^2^ increase in BMI.

Patients were randomized within each trial to receive the pan-PPAR agonist chiglitazar or a trial-specific comparator (**Fig. 1D**). The 350 patients from the placebo-controlled trial (CMAP, NCT02121717, ClinicalTrials.gov)^85^ were randomly assigned to receive low-dose chiglitazar (32 mg/day, *n* = 106), high-dose chiglitazar (48 mg/day, *n* = 106), or placebo (*n* = 138). The 485 patients from the sitagliptin-controlled trial (CMAS, NCT02173457, ClinicalTrials.gov)^86^ were randomly assigned to receive low-dose chiglitazar (32 mg/day, *n* = 161), high-dose chiglitazar (48 mg/day, *n* = 161), or sitagliptin (*n* = 163). Baseline clinical characteristics were largely comparable among treatment groups within each trial and between the two trials (**Table S1**).

Patients showed heterogeneity in homeostatic model assessment of IR (HOMA-IR) levels at baseline, ranging from 0.63 to 53.1 (**Fig. 1E**). We defined IR individuals with HOMA-IR thresholds of ≥ 4.31 for males and ≥ 4.51 for females, based on criteria established for the Chinese population^88^. Among the 826 drug-naïve patients with available baseline HOMA-IR data, 409 (49.5%) were categorized as IR (**Fig. 1E**). IR was closely coupled with obesity: IR individuals showed a significantly higher prevalence of obesity (*n* = 314 [76.8%] vs. 195 [46.8%], Fisher’s exact *P* < 0.0001) (**Fig. 1F**) and higher BMI (27.0 [25.1, 28.9] vs. 24.7 [23.1, 26.6] kg/m^2^, Wilcoxon *P* < 0.0001) than non-IR individuals (**Fig. 1G**). However, 37 of 409 IR individuals were normal weight (**Figs. 1F-G**), showing that diabetes-related IR can occur without obesity.

Chiglitazar reversed IR in about half of patients, compared with fewer than a third on placebo or sitagliptin. Among 358 participants with baseline IR and available week-24 assessments, IR-reversal proportions for low-dose chiglitazar, high-dose chiglitazar, and the trial-specific comparator were 55.8%, 45.1%, and 28.6% in CMAP and 51.5%, 59.2%, and 31.5% in CMAS, respectively (**Fig. 1H**). Pooled across doses and trials, 53.4% (126/236) of chiglitazar-treated patients reversed IR, compared with 28.6% (14/49) on placebo and 31.5% (23/73) on sitagliptin. Reversal was significantly more frequent than with the comparator for both doses in CMAS and for the low dose in CMAP, but not for the high dose in CMAP (Holm-adjusted *P* = 0.202) (**Fig. 1H**). In all participants, trial-specific mixed-effects models estimated week-24 HOMA-IR changes of −27.5 ± 3.6%, −35.2 ± 3.2% and −5.9 ± 4.4% in CMAP and −24.3 ± 3.3%, −29.7 ± 2.9% and −5.1 ± 3.9% in CMAS, in the same treatment order (estimate ± standard error [SE]). Both doses produced greater reductions than their respective controls (all Holm-adjusted dose-control *P* < 0.001; **Fig. 1I**).

Chiglitazar improved insulin sensitivity despite concurrent BMI increases. At week 24, estimated BMI changes were 2.1 ± 0.3%, 2.7 ± 0.4% and −0.7 ± 0.3% in CMAP and 1.3 ± 0.3%, 1.7 ± 0.3% and −0.1 ± 0.2% in CMAS for low-dose chiglitazar, high-dose chiglitazar and the trial-specific comparator, respectively (all Holm-adjusted dose-control *P* < 0.001; **Fig. 1J**). HOMA-IR changes were positively associated with BMI changes in the placebo and sitagliptin arms (*β* = 0.184, Holm-adjusted *P* = 0.0032 and *β* = 0.253, Holm-adjusted *P* = 2.45 × 10^−5^, respectively). In contrast, the corresponding associations with low- or high-dose chiglitazar were not significant (**Fig. 1K**). Thus, pan-PPAR agonism moved insulin sensitivity and BMI in opposite directions, uncoupling insulin sensitization from weight loss.

We generated a longitudinal multi-omics dataset of over 2,000 serial fasting blood samples from the ChiHOPE cohort (**Fig. S1A-B**). We implemented stringent quality control (QC) procedures to ensure data quality^89–94^ (**Figs. S1C-G**, **Methods**). To separate IR molecular signatures from those of body weight, we integrated metabolomic and proteomic data into molecular HOMA-IR scores and traced their changes during pan-PPAR agonism. We then tested whether molecular IR classifications distinguished metabolic heterogeneity within conventional IR categories. Finally, we decomposed molecular contributions to IR reversal and investigated sex-associated differences in the response to chiglitazar.

### A plasma multi-omics estimate of HOMA-IR outperforms routine clinical measures

Leveraging baseline plasma molecular measurements (217 metabolites and 837 proteins), we trained elastic-net regression models to predict baseline HOMA-IR in 705 ChiHOPE patients with paired metabolomics, proteomics, and clinical measures. Key baseline glycemic characteristics, including HbA1c and measured HOMA-IR, were similar between included and excluded participants (**Table S2**). We fitted separate elastic-net models for metabolomics and proteomics and combined their training-set cross-fitted predictions using OLS regression to produce metabolomics-based HOMA-IR (met-HOMA-IR), proteomics-based HOMA-IR (prot-HOMA-IR), and integrated molecular HOMA-IR (mol-HOMA-IR) models, collectively termed molecular HOMA-IR estimates. We developed the models using nested cross-validation with 20 repetitions of ten-fold outer cross-validation, and averaged the hold-out predictions to obtain a single estimate for each patient (**Figs. 2A** and **S2**).

**Figure 2.**
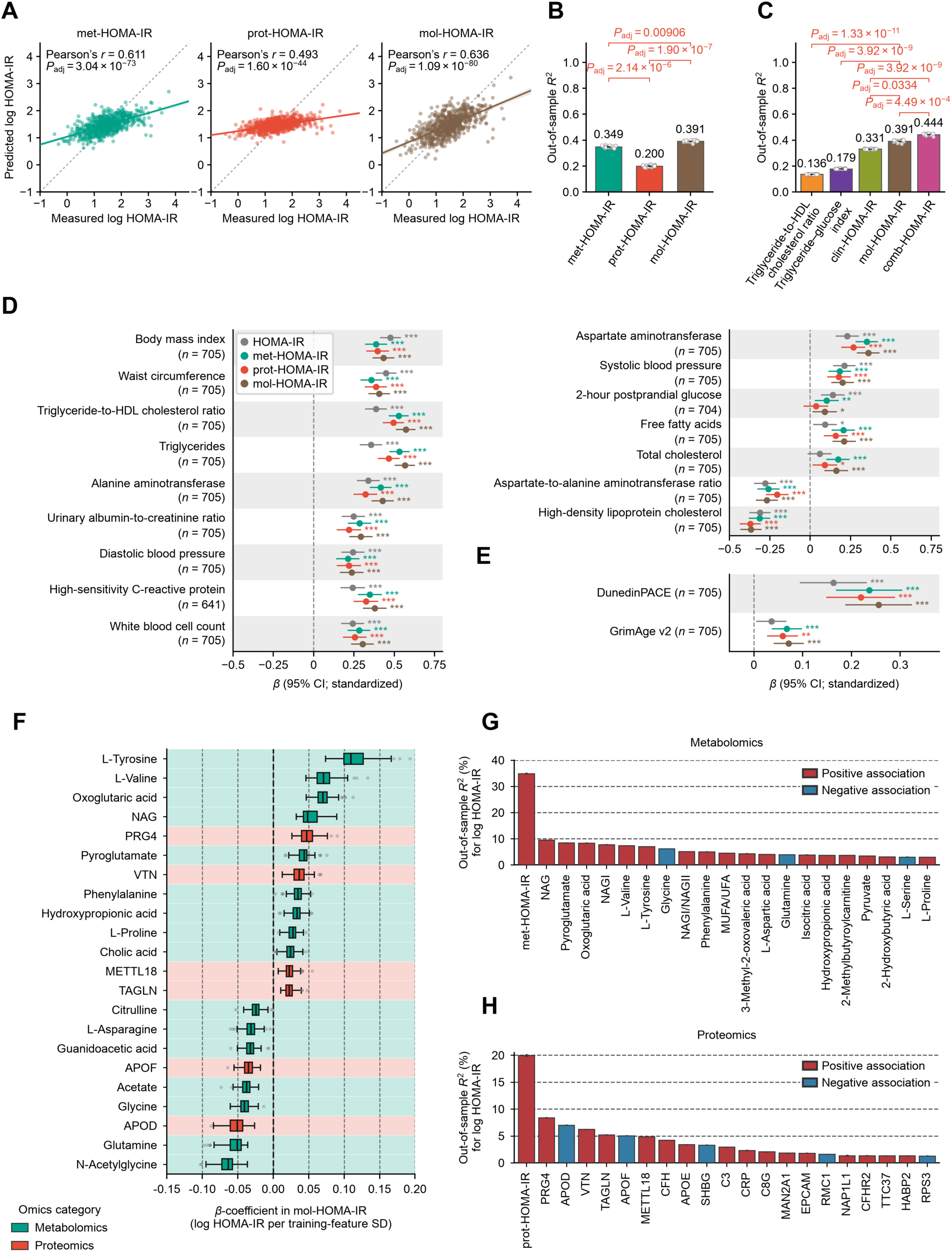
A plasma multi-omics estimate of HOMA-IR outperforms routine clinical measures. (**A**) Correlation between the measured and predicted log-transformed HOMA-IR in 705 participants at baseline. The solid line is the OLS linear regression line with a 95% confidence interval, and the dotted line is measured log-transformed HOMA-IR = predicted log-transformed HOMA-IR. *P*_adj_: BH-adjusted *P* value of a two-sided Pearson’s correlation test across the three categories. (**B-C**) Model performance of each fitted HOMA-IR model. For each repetition, out-of-sample *R*^2^ was calculated by pooling held-out predictions across all ten outer folds. Bars show mean ± standard error (SE) across 20 repetitions. Numbers above bars indicate mean *R*^2^. *P*_adj_: BH-adjusted *P* value of a two-sided Nadeau–Bengio corrected paired t-test on MSE differences across the 200 outer test folds. BH correction covered three pairwise comparisons in B and all ten pairwise comparisons in C; five selected comparisons are displayed in C. (**D-E**) Association between molecular HOMA-IR estimates and clinical measures (**D**) and epigenetic aging measures (**E**). For each of the 18 clinical measures and 11 epigenetic aging measures, *a β*-coefficient was estimated using an OLS linear regression model with each measure as the dependent variable and the measured or molecular HOMA-IR estimate as the predictor. Models were adjusted for sex, age and clinical trial, and models for epigenetic aging measures were additionally adjusted for estimated blood-cell composition. Analyses used available observations from the 705-participant baseline cohort. HOMA-IR predictors and clinical outcomes were analyzed on the natural-log scale and standardized; epigenetic aging outcomes were analyzed on their original scales, with coefficients divided by the outcome SD for display. Multiple testing correction was performed using the BH method across 72 clinical regressions (18 measures × four HOMA-IR types) and 44 epigenetic aging regressions (11 measures × four HOMA-IR types). *n*, number of assessed participants. Data: standardized *β*-coefficients with 95% confidence intervals. *adjusted *P* < 0.05, **adjusted *P* < 0.01, ***adjusted *P* < 0.001. All exact values of test summaries are found in Source Data. (**F**) Coefficients of the 22 analytes (16 metabolites and six proteins) retained in all 200 outer-fold models (20 repetitions × ten folds) in 705 participants. Coefficients incorporate fold-specific integration weights and represent log HOMA-IR per standard deviation of each analyte within each training fold. Analytes are ordered by decreasing median signed *β*-coefficient. Background and box colors indicate the omics category. Boxes show the interquartile range, central lines indicate medians, whiskers extend to the most extreme observations within 1.5 times the interquartile range, and points indicate outliers. (**G, H**) Out-of-sample predictive performance of individual metabolites (**G**) and proteins (**H**) for log HOMA-IR. Single-analyte OLS models were evaluated for the 83 metabolites and 65 proteins selected in at least one of the ten outer-fold models in every cross-validation repetition. Each model included an intercept and one analyte, without additional covariates, and used the same 705 participants and outer-fold assignments as the corresponding full omics model. The 20 analytes with the highest mean out-of-sample *R*^2^ are shown alongside the full metabolomics or proteomics model as a reference. Bars show mean *R*^2^ across 20 repetitions, expressed as a percentage; error bars indicate the SE across repetitions. Red and blue indicate positive and negative mean single-analyte regression coefficients, respectively; full-model reference bars are shown in red.

Multi-omics integration explained more variance in HOMA-IR than either single-omics platform. The mol-HOMA-IR model significantly outperformed met-HOMA-IR and prot-HOMA-IR (mean out-of-sample *R*^2^ = 0.391, 0.349, 0.200, respectively; **Fig. 2B**), supporting complementary IR information across the two omics platforms. The mol-HOMA-IR model significantly outperformed routine clinical surrogates^95–97^: mean out-of-sample *R*^2^ for HOMA-IR was 0.136 for the triglyceride-to-HDL cholesterol ratio^95^, 0.179 for the triglyceride-glucose (TyG) index^96^, and 0.331 for a clinical model including BMI, waist circumference, triglycerides, HDL cholesterol and ALT (clin-HOMA-IR), rising to 0.444 when mol-HOMA-IR was added (comb-HOMA-IR) (**Fig. 2C**), indicating that plasma multi-omics provides IR information beyond traditional clinical measurements. This advantage in mol-HOMA-IR was maintained in 593 individuals at week 24 (**Fig. S3A-C**).

HOMA-IR was significantly associated with obesity (BMI, waist circumference), lipid measures (triglycerides, HDL cholesterol and free fatty acids), liver- and kidney-risk measures (alanine aminotransferase [ALT], aspartate aminotransferase [AST], AST-to-ALT ratio^98^, and urinary albumin-to-creatinine ratio [UACR]), blood pressure (diastolic and systolic blood pressure), systemic inflammation measures (high-sensitivity C-reactive protein [hsCRP] and white blood cell count [WBC]) and an epigenetic measure of the pace of aging (DunedinPACE)^99^ (**Figs. 2D-E, Fig. S3D**). Each molecular HOMA-IR estimate showed concordant associations in the same direction as measured HOMA-IR, with comparable effect sizes for these measures (**Figs. 2D-E, Fig. S3D**). This concordance indicates that the molecular estimates retained the clinical profile of measured HOMA-IR. To test the generalizability of our results, we examined 333 participants from NSPT^100^, an independent cohort of Han Chinese volunteers from Zhengzhou, China, with plasma metabolomics, proteomics and clinical measures but without HOMA-IR. We calculated HOMA-IR predictions for the NSPT cohort with restricted models that were fitted to the ChiHOPE datasets (**Fig. S4A-B**). In the NSPT cohort, molecular HOMA-IR estimates were also associated with obesity, lipid measures, liver-risk measures and blood pressure (**Fig. S4C**). The direction and magnitude of these associations were concordant with those in ChiHOPE, supporting the transferability of the clinical relevance of molecular HOMA-IR estimates.

We characterized the molecular signature underlying molecular HOMA-IR estimates. Among the 148 analytes (83 metabolites and 65 proteins) that were selected in at least one of the ten outer-fold models in every cross-validation repetition, 16 metabolites and six proteins were retained in all outer-fold models (**Fig. S5A**). The most stable and strongest predictors in the mol-HOMA-IR model were tyrosine, valine, oxoglutaric acid, N-acetylated glycoproteins (NAG)^101,102^, and proteoglycan 4 (PRG4) as positive predictors, and N-acetylglycine, glutamine, and apolipoprotein D (APOD) as negative predictors (**Fig. 2F**). We then tested each of these analytes individually. The strongest individual metabolomic and protein predictors, NAG and PRG4, explained 9.5% and 8.4% of the variance in HOMA-IR, respectively, substantially less than the full metabolomic and proteomic models (34.9% and 20.0%, respectively; **Fig. 2G-H**). Moreover, after sequential removal of leading predictors including NAG and PRG4, the models still explained more variance in HOMA-IR than any single analyte (**Fig. S5B-D**). Thus, molecular HOMA-IR reflects coordinated changes across multiple analytes on both platforms, rather than a single dominant marker.

### Molecular IR reveals metabolic risk hidden within HOMA-IR classes

Although mol-HOMA-IR explained 39.1% of the variance in classically measured HOMA-IR, molecular estimates differed from measured values (**Fig. 2A**). To quantify this difference, we defined molecular HOMA-IR gaps as residual deviations of molecular estimates from their expected values given measured HOMA-IR. The metabolomic and proteomic gaps, which derive from independent assays, were correlated (Pearson’s *r* = 0.398, *P* < 0.0001), suggesting that the deviations reflect a shared physiological signal rather than assay noise. These gaps were significantly associated with lipid measures (triglycerides, HDL cholesterol and free fatty acids), liver- and kidney-risk measures (ALT, AST, AST-to-ALT ratio and UACR), systemic inflammation measures (hsCRP and WBC), blood pressure, and epigenetic aging measurements (DunedinPACE^99^ and GrimAge v2^103^) (**Fig. 3A-B**), indicating molecular HOMA-IR estimates captured clinical and aging heterogeneity beyond measured HOMA-IR.

**Figure 3.**
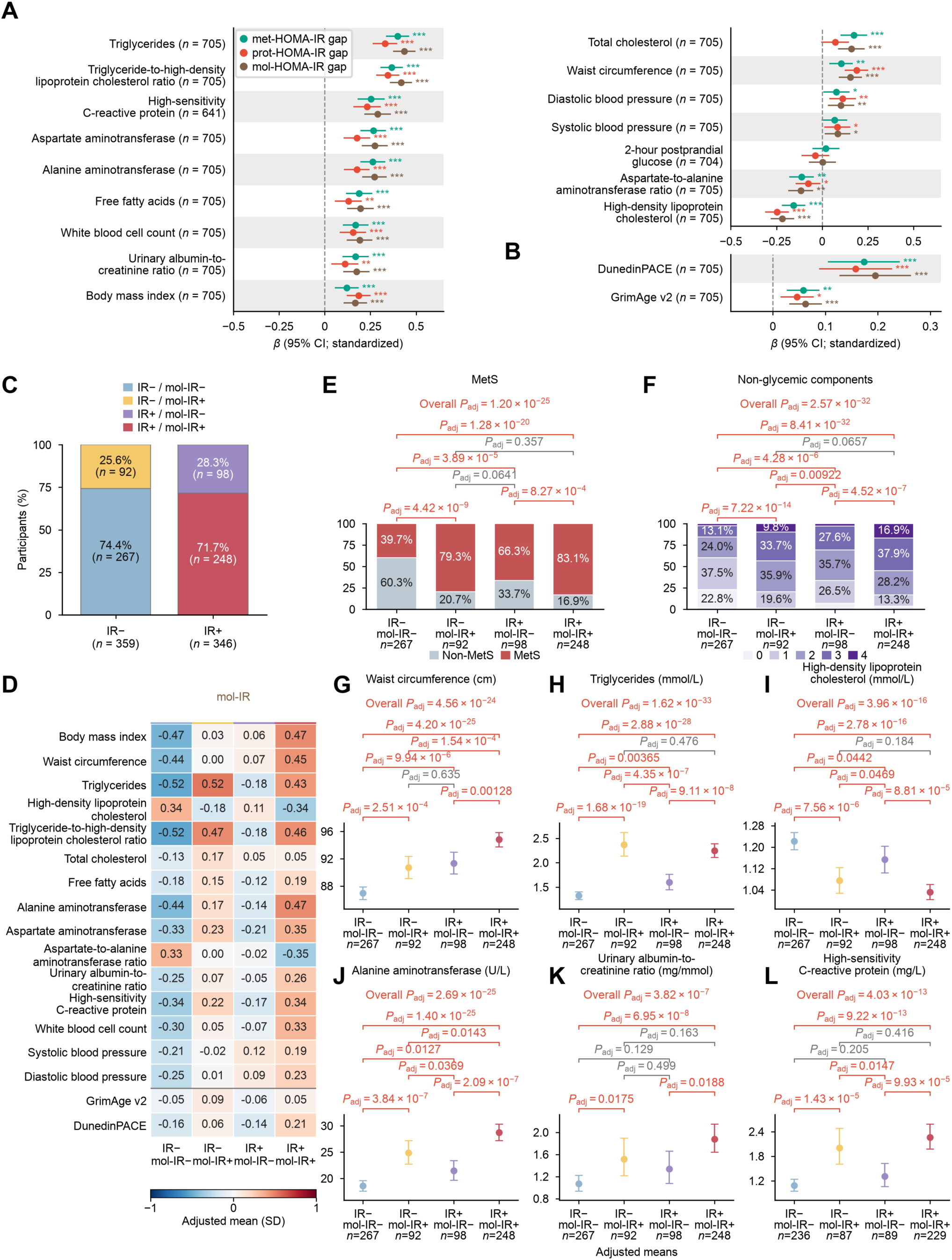
Molecular IR reveals metabolic risk hidden within HOMA-IR classes. Analyses used available observations from the 705-participant baseline cohort. (**A**-**B**) Association between molecular HOMA-IR gaps and clinical measures (**A**) and epigenetic aging measures (**B**). Gaps were calculated as residual deviations of predicted log-transformed HOMA-IR from the value expected given measured log-transformed HOMA-IR, using calibration models fitted within the outer training sets, and averaged across 20 cross-validation repetitions. For each of the 18 clinical measures and 11 epigenetic aging measures, *a β*-coefficient was estimated using an OLS linear regression model with each measure as the dependent variable and each gap as the predictor. Models were adjusted for log-transformed HOMA-IR, sex, age and clinical trial, and models for epigenetic aging measures were additionally adjusted for estimated blood-cell composition. Multiple testing correction was performed using the BH method across 54 clinical regressions (18 measures × three gaps) and 33 epigenetic aging regressions (11 measures × three gaps). *n*, number of assessed participants. Data: estimates with 95% confidence intervals. *adjusted *P* < 0.05, **adjusted *P* < 0.01, ***adjusted *P* < 0.001. All exact values of test summaries are found in Source Data. (**C**) Distribution of mol-IR status within measured IR− and IR+ groups. Measured and molecular IR were defined using sex-specific HOMA-IR thresholds of ≥4.31 in men and ≥4.51 in women. (**D**) Covariate-adjusted means of 15 clinical measures and two epigenetic aging measures across the four groups, expressed in standard-deviation units. We log-transformed the clinical measures before standardization. (**E**-**F**) Observed distributions of metabolic syndrome (MetS; **E**) and the number of non-glycemic MetS components (**F**). The four non-glycemic components were abdominal obesity, elevated blood pressure, elevated triglycerides and low HDL cholesterol. (**G**-**L**) Adjusted geometric means and 95% confidence intervals for waist circumference (**G**), triglycerides (**H**), HDL cholesterol (**I**), alanine aminotransferase (**J**), urinary albumin-to-creatinine ratio (**K**) and high-sensitivity C-reactive protein (**L**). Four-group models adjusted for sex, age and clinical trial, with additional blood-cell adjustment for epigenetic aging measures. Adjusted means were standardized to the covariate distribution of the eligible analysis sample. Overall and pairwise differences were assessed using OLS F- and t-tests for continuous measures, logistic likelihood-ratio and Wald tests for MetS, and OLS with HC3 robust F- and t-tests for component counts. For continuous clinical measures, BH adjustment covered 54 overall tests and 324 pairwise comparisons across three molecular classifications. For MetS-related outcomes, adjustment covered 18 overall tests and 108 pairwise comparisons across six outcomes and three classifications.

We defined met-IR, prot-IR, and mol-IR using the same thresholds as for standard IR classes. At baseline, measured IR and mol-IR classifications were discordant in 27% of participants (190/705), yielding four groups: IR−/mol-IR− (*n* = 267), IR−/mol-IR+ (*n* = 92), IR+/mol-IR− (*n* = 98) and IR+/mol-IR+ (*n* = 248) (**Fig. 3C**). Within each measured IR class, mol-IR+ participants generally exhibited less favorable metabolic profiles than mol-IR− participants (**Fig. 3D**), with a higher prevalence of metabolic syndrome (MetS) and its non-glycemic components (**Fig. 3E-F**), especially abdominal obesity (**Fig. 3G**), elevated triglycerides (**Fig. 3H**) and low HDL cholesterol (**Fig. 3I**). Also, mol-IR+ participants showed higher liver- and kidney-risk measures (**Figs. 3D**, **3J-K**), and systemic inflammation measures (**Figs. 3D** and **3L**). Classifications based on met-HOMA-IR and prot-HOMA-IR showed the same pattern (**Fig. S6**). Together, these findings suggest that IR classification based on molecular estimates captured metabolic heterogeneity within conventional IR categories.

Molecular HOMA-IR estimates retained associations with obesity: both the estimates and their residual gaps were significantly associated with BMI and waist circumference (**Figs. 2D** and **3A**). Within each measured IR category, mol-IR+ participants generally had higher BMI and waist circumference than mol-IR− participants (**Fig. 3D**). To assess whether weight accounted for the clinical and epigenetic associations, we additionally adjusted for both BMI and waist circumference. Associations between molecular HOMA-IR gaps and lipid, liver, and inflammatory measures, as well as GrimAge v2 and DunedinPACE, remained significant (**Fig. S7A-B**). Within both measured IR categories, mol-IR+ participants continued to exhibit higher triglycerides, ALT and hsCRP and lower HDL cholesterol than mol-IR− participants after adjustment (**Fig. S7C-K**). Clinical associations were also retained in sensitivity models including BMI as an unpenalized covariate (**Fig. S8**). These findings suggest that molecular HOMA-IR captures clinical and epigenetic variation not fully accounted for by measured HOMA-IR and body weight.

### IR-linked molecules improve as body weight rises

Because IR and body weight always covary cross-sectionally, we next used the randomized pan-PPAR perturbation, which reverses IR despite weight gain, to separate their molecular signatures. These analyses included 593 participants from the 705-participant baseline cohort with paired baseline and week-24 metabolomic and proteomic measures (CMAP, *n* = 219; CMAS, *n* = 374). Baseline HbA1c and measured HOMA-IR were similar between participants included in these analyses and those excluded from the original cohort, and inverse probability weighting yielded comparable treatment-effect estimates (**Table S3**). At baseline, mol-HOMA-IR estimates and mol-IR classifications did not differ significantly among treatment groups within either trial (**Fig. S9**). By week 24, both chiglitazar doses produced greater adjusted reductions in molecular HOMA-IR estimates than their respective comparators (**Fig. 4A-B**), resulting in a larger proportion of participants without mol-IR at week 24 (**Fig. 4C-D**). These reductions occurred despite increases in mean BMI in both chiglitazar arms of each trial (**Fig. 1J**), indicating that molecular IR can improve despite weight gain.

**Figure 4.**
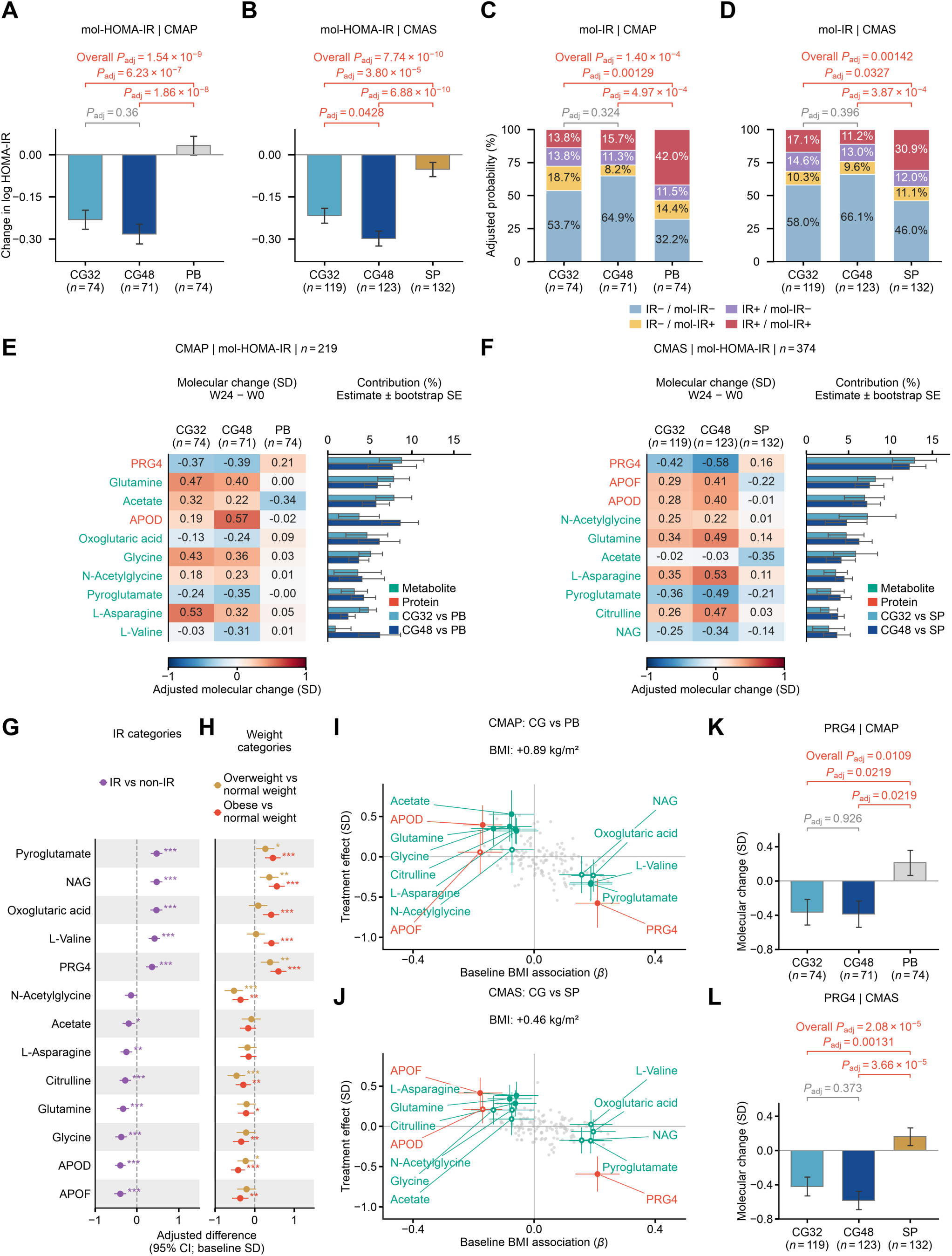
IR-linked molecules improve as weight rises. (**A-B**) Adjusted changes in mol-HOMA-IR from baseline to week 24 in CMAP (**A**) and CMAS (**B**). Analyses included participants from the 705-participant baseline cohort with paired measurements and complete non-genetic covariates (CMAP, *n* = 219; CMAS, *n* = 374). Adjusted changes were estimated using ANCOVA with week-24 mol-HOMA-IR as the dependent variable and treatment group, baseline mol-HOMA-IR, sex, age as predictors. Molecular HOMA-IR estimates are already expressed on the natural-log HOMA-IR scale and were not log-transformed again. Data: adjusted mean change ± s.e. Overall and pairwise differences were assessed using partial *F*-tests and two-sided *t*-tests. (**C-D**) Adjusted week-24 probabilities of the four categories defined by measured IR and mol-IR status in CMAP (**C**) and CMAS (**D**). We estimated probabilities using mean-bias-reduced multinomial logistic regression, adjusting for baseline four-category status, sex and age, and standardized the probabilities to the covariate distribution of the eligible analysis sample. Analyses included all eligible paired participants, irrespective of baseline IR status. Measured and molecular IR were defined using sex-specific HOMA-IR thresholds of ≥4.31 in men and ≥4.51 in women. Overall and pairwise differences were assessed using joint Wald tests. For A–D, BH adjustment covered seven overall tests and 21 pairwise comparisons within each trial across four continuous HOMA-IR measures and three molecular classifications. (**E-F**) Adjusted molecular changes and contributions to treatment-associated reductions in mol-HOMA-IR in CMAP (**E**) and CMAS (**F**). Candidate analytes comprised 83 metabolites and 65 proteins selected in at least one of the ten outer-fold models in every cross-validation repetition. Within each trial, analytes contributing to lower mol-HOMA-IR in both dose–control comparisons were ranked by their mean contribution across the two doses; the top ten are presented. Heatmaps show adjusted molecular changes from baseline to week 24, expressed in frozen baseline-training SD units. Contribution bars show each analyte’s percentage of the summed lowering-direction contributions across all analytes, including those not displayed. Models adjusted for baseline mol-HOMA-IR, sex, age. Error bars represent s.e. estimated from 2,000 participant bootstrap resamples within trial and treatment arm, with molecular models held fixed. Green and red labels indicate metabolites and proteins, respectively. (**G-H**) Baseline associations of the 13 unique analytes presented in E–F with measured IR (**G**) and weight categories (**H**) among participants with complete covariates (*n* = 705). For each analyte, *β*-coefficients were estimated using separate OLS linear regression models with standardized molecular levels as the dependent variable and IR status or weight category as the predictor. Models were adjusted for sex, age, clinical trial. Normal weight, overweight and obesity were defined as BMI 18.5–<23, 23–<25 and ≥25 kg/m², respectively. Data: adjusted difference with 95% confidence interval. Two-sided *t*-tests were used, retaining BH adjustment across the original 51 comparisons (17 analytes × three contrasts). We retained all 13 displayed analytes, regardless of statistical significance. (**I-J**) Baseline associations with BMI plotted against randomized chiglitazar effects in CMAP (**I**) and CMAS (**J**). The horizontal axis shows *β*-coefficients from OLS models with standardized molecular levels as the dependent variable and standardized BMI as the predictor, adjusting for sex, age, clinical trial. The vertical axis shows the equally weighted average of the two chiglitazar dose–control contrasts, estimated using ANCOVA with week-24 molecular levels as the dependent variable, adjusting for the corresponding baseline molecular level, sex, age. Molecular effects are expressed in baseline SD units. Grey points represent all 148 candidate analytes; the 13 analytes shown in G–H are highlighted and labeled. Horizontal and vertical error bars indicate 95% confidence intervals. Filled and hollow markers indicate treatment BH-adjusted *P* < 0.05 and ≥0.05, respectively, with adjustment across 148 analytes within each trial and contrast. Corresponding adjusted BMI treatment effects are annotated. (**K-L**) Adjusted changes in PRG4 from baseline to week 24 in CMAP (**K**) and CMAS (**L**), using the same standardization and covariate adjustment as in **E**-**F**. Data: adjusted mean change ± s.e. Overall and pairwise differences were assessed using partial *F*-tests and two-sided *t*-tests. BH adjustment covered 13 overall tests and 39 pairwise comparisons within each trial. Treatment-effect models were not adjusted for BMI change. *n*, number of assessed participants; *P*_adj_, BH-adjusted *P* value. *adjusted *P* < 0.05, **adjusted *P* < 0.01, ***adjusted *P* < 0.001. CG32 and CG48, chiglitazar treatment groups; PB, placebo; SP, sitagliptin.

Decomposition of the chiglitazar-control differences in mol-HOMA-IR reductions identified the molecular features that contributed most. Among the 148 analytes that were selected in at least one of the ten outer-fold models in every cross-validation repetition, 85 analytes (51 metabolites and 34 proteins) contributed to lower mol-HOMA-IR with both chiglitazar doses relative to placebo in the CMAP trial, led by PRG4, glutamine, acetate and APOD (**Fig. 4E**). Meanwhile, 80 analytes (42 metabolites and 38 proteins) contributed to lower mol-HOMA-IR with both chiglitazar doses relative to sitagliptin in the CMAS trial, led by PRG4, APOF, APOD and N-acetylglycine (**Fig. 4F**). A total of 52 analytes (31 metabolites and 21 proteins) contributed to lower mol-HOMA-IR with both chiglitazar doses relative to the respective control in both trials, representing a candidate set of IR-linked molecules that pan-PPAR agonism moved despite BMI gain. These molecular signatures included amino acid metabolites, such as glycine and pyroglutamate, and lipid-transport proteins, including APOD and APOF (**Fig. S10A**). Acetate rose with chiglitazar in CMAP and was stable in CMAS, whereas it fell with both placebo and sitagliptin, so its contribution partly reflects changes in the comparator arms. However, several features with substantial weights in the mol-HOMA-IR model, including tyrosine and phenylalanine, did not contribute in the same directions across the two dose-control contrasts within each trial.

Cross-sectionally, 18 of these 52 signatures (34.6%) were significantly associated with both IR and BMI (**Fig. S10A**), so at baseline their relationship with IR could not be separated from body weight. During chiglitazar treatment, however, features linked to larger BMI moved in the insulin-sensitizing direction while BMI rose (**Fig. 4I-J**). Changes in 51 of the 52 features were not significantly associated with changes in BMI (**Fig. S10B**), and 34 of the 41 significant baseline associations with IR persisted after adjustment for BMI (**Fig. S10C**). As the leading contributor in both trials, PRG4 decreased significantly compared with placebo and sitagliptin (**Figs. 4K-L**) and contributed to lower mol-HOMA-IR relative to the respective control despite BMI gain (**Figs. 4E-F**). PRG4 was associated with both BMI and IR at baseline (**Figs. 4G-H**, **Fig. S10A**). Thus, randomized pan-PPAR agonism revealed changes in IR-associated molecular features despite BMI gain, complementing their cross-sectional associations with BMI.

### Molecular improvement tracks clinical benefit beyond measured HOMA-IR

In the parent trials^85,86^, chiglitazar improved glycemic control and multiple clinical measures. Among chiglitazar-treated participants, reductions in mol-HOMA-IR were associated with clinical improvement after adjustment for changes in measured HOMA-IR, including decreases in 2-hour glucose (measured from meal test), triglycerides, free fatty acids, ALT, hsCRP and WBC, as well as increases in HDL cholesterol (**Fig. 5A**). Concordantly, mol-IR− status at week 24 was associated with more favorable clinical changes within measured IR categories. For example, mol-IR− participants showed greater triglyceride reductions in both measured IR categories and, within the IR+ category, greater reductions in 2-hour glucose and ALT (**Figs. 5B-E**). Together, these findings link molecular remodeling to metabolic improvement during pan-PPAR agonism.

**Figure 5.**
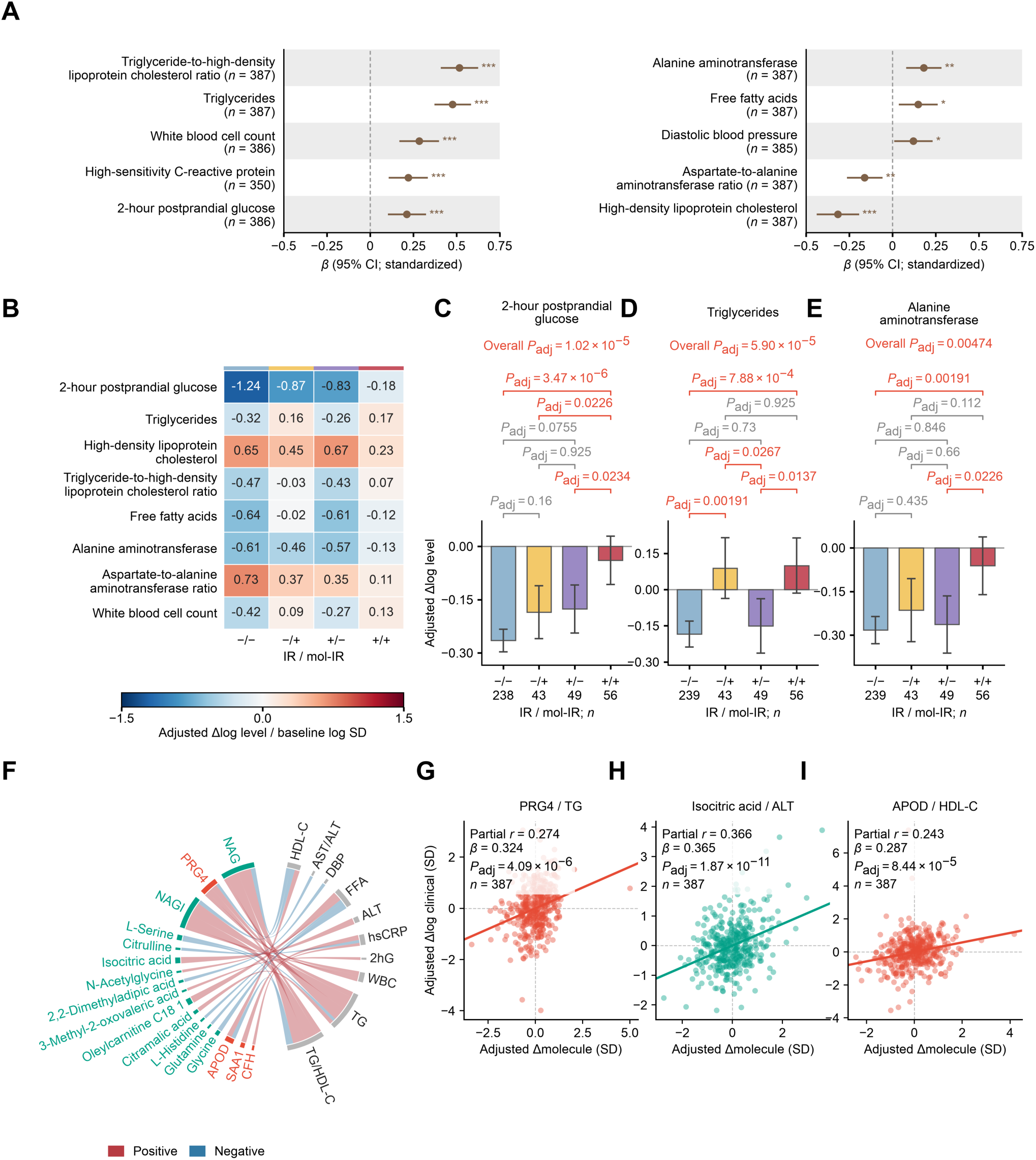
Molecular improvement tracks clinical benefit beyond measured HOMA-IR. Analyses used molecular HOMA-IR estimates derived from the models developed in the 705-participant baseline cohort and included 387 participants receiving chiglitazar 32 or 48 mg with paired baseline and week-24 data and complete non-genetic baseline covariates (CMAP, *n* = 145; CMAS, *n* = 242). Sample sizes varied according to outcome and analyte availability. (**A**) Associations between changes in mol-HOMA-IR and clinical measures. For each clinical measure, we regressed the standardized log change on the standardized change in mol-HOMA-IR using OLS linear regression. Models were adjusted for baseline log clinical level, baseline mol-HOMA-IR, baseline measured log HOMA-IR, change in measured log HOMA-IR, sex, age, clinical trial, dose. Multiple testing correction was performed using the Benjamini–Hochberg (BH) method across 54 regressions (18 clinical measures × three molecular estimates). Only the ten clinical measures significantly associated with changes in mol-HOMA-IR after adjustment are presented. Data: standardized *β*-coefficient with 95% confidence interval. (**B**) Covariate-adjusted clinical changes across the four week-24 IR/mol-IR groups, expressed as log changes divided by the baseline log standard deviation. The eight displayed measures showed an overall significant difference and at least one significant pairwise difference between mol-IR− and mol-IR+ participants within the same measured IR category in either the main or sensitivity model. The sensitivity model replaced baseline four-group status with baseline measured and molecular log HOMA-IR as continuous covariates; the heatmap presents estimates from the main model. (**C**–**E**) Adjusted log changes and 95% confidence intervals for 2-hour postprandial glucose (C), triglycerides (D) and alanine aminotransferase (E). Measured and molecular IR were defined using sex-specific HOMA-IR thresholds of ≥4.31 in men and ≥4.51 in women. Four-group models were adjusted for baseline log clinical level, baseline four-group status, sex, age, clinical trial, dose. Adjusted means were standardized to the covariate distribution of the eligible sample for each outcome. Overall and pairwise differences were assessed using OLS F-tests and two-sided t-tests. BH adjustment covered 18 overall tests and 108 pairwise comparisons, separately for each model. Red pairwise annotations indicate comparisons meeting both the overall and pairwise adjusted *P* < 0.05 criteria; other annotations are gray. (F) Associations between changes in 52 analytes and 11 clinical measures (the ten measures displayed in A plus HbA1c). The analytes included 31 metabolites and 21 proteins that contributed to lower mol-HOMA-IR with both chiglitazar doses relative to the respective control in both trials. Standardized clinical log changes were regressed on standardized analyte changes, adjusting for baseline analyte level, baseline log clinical level, baseline measured log HOMA-IR, change in measured log HOMA-IR, sex, age, clinical trial, dose. BH correction covered all 572 regressions (52 analytes × 11 clinical measures). The chord diagram presents 28 significant associations involving 17 analytes and ten clinical measures. Red and blue ribbons indicate positive and negative associations, respectively; ribbon widths represent absolute standardized *β*-coefficients. Molecular labels are colored by omics category. Only analytes and clinical measures with significant associations are displayed. (G–I) Covariate-adjusted associations between changes in PRG4 and triglycerides (G), isocitric acid and alanine aminotransferase (H), and APOD and HDL cholesterol (I). Both plotted variables were residualized against the covariates specified for F. Points represent individual participants, and lines represent fitted linear associations. Partial *r*, standardized *β* and BH-adjusted *P* values correspond to the models in F. Clinical changes were calculated as ln(week 24) − ln(baseline); analyte changes were calculated from the existing transformed model inputs. Models did not additionally adjust for BMI change. *n*, number of assessed participants. *Adjusted *P* < 0.05, **adjusted *P* < 0.01, ***adjusted *P* < 0.001.

We next examined how changes in each molecular signature were associated with metabolic improvement. Among the 52 analytes previously identified as contributing to lower mol-HOMA-IR with both chiglitazar doses relative to the respective control in both trials, we identified 28 significant associations with week-24 changes in 11 clinical measures (**Fig. 5F**). For example, changes in PRG4 were positively associated with changes in triglycerides, changes in isocitric acid with changes in ALT, and changes in APOD with changes in HDL cholesterol (**Figs. 5G-I**). These findings support coordinated molecular and clinical changes beyond their shared association with changes in measured HOMA-IR.

### Women showed greater molecular IR reversal with pan-PPAR agonism

Molecular IR responses to pan-PPAR agonism were heterogeneous. Among 387 chiglitazar-treated participants with paired baseline and week-24 classifications, 88.0% (125/142) of those classified as IR−/mol-IR− at baseline remained in this category at week 24. Among participants classified as IR+/mol-IR− or IR−/mol-IR+ at baseline, 65.3% (32/49) and 55.0% (33/60), respectively, transitioned to IR−/mol-IR−. By comparison, only 36.0% (49/136) of those with baseline IR+/mol-IR+ status transitioned to IR−/mol-IR− (**Fig. S11A**). Among these 136 baseline IR+/mol-IR+ participants (85 men and 51 women), women were more likely than men to transition to IR−/mol-IR− and showed a greater reduction in mol-HOMA-IR estimates. Both associations were significant after adjustment for baseline measured and molecular HOMA-IR, trial and dose (**Fig. S11B-C**) and persisted after additional adjustment for age and baseline HbA1c, HDL cholesterol and triglycerides. In trial-specific analyses, women showed greater mol-HOMA-IR reductions than men in the CG32 groups of both CMAP and CMAS (**Fig. S11C**). Sex-by-treatment interactions for mol-HOMA-IR change were significant after BH correction in CMAS (adjusted *P* = 0.0127), but not in CMAP (adjusted *P* = 0.0773); interactions for the four-category outcome were not significant in either trial (CMAS, adjusted *P* = 0.185; CMAP, adjusted *P* = 0.463). The CMAP CG48 group included only five women, limiting the precision of its sex-specific estimates. These exploratory findings suggest sex-related heterogeneity in molecular IR improvement among participants with baseline IR+/mol-IR+ status, with a female advantage most consistently observed at the low dose.

## Discussion

Insulin resistance does not have to wait for weight loss. In two randomized trials, chiglitazar reversed IR in about half of patients with T2D while their BMI rose, and the molecular features that track IR moved with insulin action, largely independently of weight. Because IR and weight rise and fall together in almost every other setting, this perturbation let us see which molecular changes belong to insulin action itself. The result supports restoring insulin action as a therapeutic goal in its own right, complementing weight-centered management^17,18,44^.

The association of IR with body weight was evident at baseline, but IR improvement occurred despite BMI gain under treatment. Cross-sectionally, about a third of the shared treatment-responsive features, including PRG4, were associated with both IR and BMI, and molecular HOMA-IR gaps still tracked BMI and waist circumference. Under chiglitazar, however, these features moved in the insulin-sensitizing direction while BMI increased. The leading features linked amino acid metabolism and glycoprotein signals to liver enzymes, inflammatory markers, and lipoproteins. These findings are compatible with the multisystem organization of IR^2,6,104^. They also emphasize a limitation of interpreting individual biomarkers in isolation: a molecule may participate in several correlated physiological processes, and a molecular contribution is a statistical summary of those relationships, not a discrete mechanism.

Why should insulin action improve while BMI increases? The likely answer lies in where lipid is stored, not how much^58,59^. PPARγ activation expands the capacity of subcutaneous adipose tissue to store lipid safely, reducing spillover to the liver and muscle^60–62^, while PPARα promotes hepatic fatty acid oxidation^105^. The molecular changes accompanying reversal fit this model: lower triglycerides, ALT, and inflammatory glycoprotein signals; higher HDL-C and APOD; and shifts in amino acid and redox metabolites, with higher glutamine and glycine and lower pyroglutamate. In this view, weight gain on pan-PPAR therapy reflects healthier lipid partitioning, and IR tracks lipid-handling quality more closely than fat quantity. This is particularly relevant to East Asian patients, who develop IR at lower BMI^47^ and in whom weight-centered treatment addresses only part of the disease. Imaging of fat distribution and liver fat would test this model directly. Adiponectin-receptor regulation and adipose FGF21 signaling provide further preclinical links between PPAR activity and insulin action^57,106^. Plasma measurements cannot, however, determine tissue origin or metabolic flux. Combining incretin and PPAR receptor agonism has recently been investigated in mice as a way to integrate weight-lowering and insulin-sensitizing actions^50^.

The molecular estimate did more than reconstruct fasting HOMA-IR. It outperformed routine clinical measures and, added to them, explained more of the variation in HOMA-IR than either alone. Molecular departures from measured HOMA-IR tracked dyslipidemia, inflammation, liver abnormalities and epigenetic aging beyond measured IR. This fits human studies linking IR to mitochondrial dysfunction and molecular aging patterns^71,107^. The discordant groups further illustrated that fasting glucose-insulin physiology and molecular metabolic burden need not rank individuals the same way. Comparable clinical associations in NSPT supported transferability of this phenotypic information, although the absence of an external HOMA-IR reference precluded diagnostic validation. The NSPT analysis could not assess associations with 2-hour postprandial glucose, HbA1c, free fatty acids, UACR, WBC or hsCRP because these measurements were unavailable, and it did not establish external validation of the epigenetic aging associations. Consequently, the scores are best viewed as research measures for molecular phenotyping and response assessment, with clinical thresholds and utility still to be established.

The longitudinal analysis showed that molecular improvement spanned multiple pathways. Glutamine, glycine, and pyroglutamate contributions were consistent with changes in amino acid and related metabolic handling, whereas APOD and glycoprotein signals implicated lipid transport and inflammatory physiology. This decomposition makes the integrated score interpretable while retaining the limitation that predictive coefficients do not establish biological causality. APOD belongs to the lipocalin family, whose functions extend to lipid transport and cardiovascular biology^108^; changes in HDL-associated protein abundance should nevertheless be distinguished from HDL function^109^.

PRG4 emerged as a candidate connecting the human observational and perturbational evidence. Its cross-sectional association with IR and its longitudinal contribution to mol-HOMA-IR improvement align with metabolic protection reported in Prg4-deficient mice^110^. However, plasma abundance, tissue origin and cell-autonomous function are distinct biological questions. Tissue-specific perturbation and rescue experiments would help distinguish a functional role from a circulating correlate of disease activity. A previous plasma proteomic study also reported a reduction in PRG4 with chiglitazar treatment^111^.

Sex differences added another dimension to response heterogeneity. Women with baseline IR+/mol-IR+ status receiving chiglitazar showed a larger adjusted decline in molecular HOMA-IR. Because this analysis began from the IR+/mol-IR+ group, regression to the mean may contribute, and sex-by-treatment interactions were significant against sitagliptin but not against placebo, where small numbers of women limited precision. The data therefore suggest a context-dependent sex difference rather than a universal female advantage across doses and comparisons. Future studies incorporating menopausal status, sex hormones, drug exposure and body-fat distribution could clarify which physiological differences explain the aggregate response.

Several limitations constrain interpretation. The molecular HOMA-IR estimates were trained on HOMA-IR, which is itself correlated with BMI, so at baseline they retain part of the BMI signal; evidence for IR improvement despite weight gain comes from the randomized longitudinal data. Longitudinal omics analyses were restricted to participants with paired samples. Measured IR reversal occurred in 28.6% of placebo recipients, so categorical transitions partly reflect regression to the mean and the within-person variability of a single fasting HOMA-IR measurement. Stratified and contribution analyses used features selected in the same cohort, so they test internal consistency rather than independent replication. This companion study involved consenting Chinese, drug-naïve trial participants and may not generalize to other populations or established multidrug treatment. HOMA-IR is a fasting surrogate and does not separately measure hepatic and peripheral insulin sensitivity as a clamp-based design could. BMI adjustment does not exclude confounding by visceral or ectopic fat. Full and restricted models were refitted in the same 705-participant development cohort using different molecular panels; external evaluation additionally required cohort-level location and scale adaptation. Repeated nested cross-validation reduced model-selection bias^112^, but follow-up predictions remained within the same cohort, and classification depended on fixed population-specific thresholds. Missing molecular or clinical observations may have selected follow-up samples, and coefficient-based contribution analyses condition on the fitted scores rather than fully propagating all training uncertainty. Finally, the aging results were cross-sectional, the sex analyses were exploratory, and long-term vascular, renal and mortality outcomes were unavailable. Look AHEAD showed that improvements in intermediate metabolic measures do not necessarily reduce cardiovascular events^113^. In addition, DNA methylation measurements were unavailable at weeks 12 and 24, precluding assessment of treatment-associated changes in epigenetic aging. Exploratory analyses that added circulating miRNAs did not materially improve HOMA-IR prediction over the combined metabolomic and proteomic model, so miRNAs were not included in the final molecular estimates.

We used BMI and body weight as measures of body size and did not measure fat mass or its distribution. PPARγ agonists can expand subcutaneous adipose tissue while reducing visceral and hepatic fat, so weight gain under chiglitazar may not reflect a less healthy adipose profile. The falls in triglycerides and ALT are consistent with this, but imaging would be needed to confirm it. HOMA-IR is a fasting index that mainly reflects hepatic insulin resistance. Our molecular estimate was trained on HOMA-IR and may also capture broader metabolic dysfunction. Validation against clamp- or OGTT-derived measures of peripheral insulin sensitivity is needed.

Translating this framework will require evaluation across assay platforms, including SomaScan and Olink proteomics and alternative metabolomic assays, along with harmonization and recalibration in broader cohorts. Large biobanks such as UK Biobank could not yet be used for external validation, because their proteomic and metabolomic platforms and panels differ from those used here. Reference-material-based ratio profiling could support such harmonization^89,93^. Organ-specific molecular measurements could further help distinguish adipose, hepatic and muscle contributions to systemic IR. Such studies would test whether the concept of molecular IR can be generalized beyond the molecular panels and study population examined here. Validation for specific clinical uses will also be needed.

Weight loss is one route to better insulin action, but not the only one. These findings show that IR-associated plasma molecular abnormalities can improve during pan-PPAR agonism despite weight gain. As incretin-based therapies make weight loss the default goal in T2D, restoring insulin action deserves attention as a target in its own right. Molecular HOMA-IR estimates complement fasting HOMA-IR and body weight by capturing heterogeneous metabolic abnormalities and their treatment-associated improvement. Independent calibration, prospective outcome studies and targeted functional experiments are the next steps toward translating this framework into clinically useful patient stratification.

## A SECONDARY AUTHOR LIST

**Human Phenome Project Scientific Consortium:**

Jun Chang^1^, Wei Chen^1^, Chen Ding^1^, Li Jin^1^, Tiemin Liu, Jingchun Luo^1^, Qiang Luo, Yuan Qi^2^, Feng Qian^1^, Leming Shi^1^, Deian Ta^1^, Huiru Tang^1^, Weishuo Tao^1^, Mei Tian^1^, Jiucun Wang^1^, Sijia Wang^3^, Shuhua Xu^1^, Zhiqiang Yan^1^, Zhong Yang^1^, Guoqing Zhang^4^, Menghan Zhang^1^, Yan Zheng^1^, and Lei Zhu^1^

**Affiliations:**

1State Key Laboratory of Genetic Engineering, School of Life Sciences, Human Phenome Institute, Fudan University, Shanghai 200433, China

^2^Artificial Intelligence Innovation and Incubation Institute, Fudan University, Shanghai 201203, China

^3^CAS Key Laboratory of Computational Biology, Shanghai Institute of Nutrition and Health, University of Chinese Academy of Sciences, Chinese Academy of Sciences, Shanghai 200031, China

^4^Bio-Med Big Data Center, Chinese Academy of Sciences Key Laboratory of Computational Biology, Shanghai Institute of Nutrition and Health, University of the Chinese Academy of Sciences, Chinese Academy of Sciences, Shanghai 200031, China

**Human Phenome Project Technological Consortium**^1^:

Ran An^1^, Wenyan Chai^1^, Chen Chen^1^, Longlong Chen^1^, Qinsheng Chen^1^, Weixin Chen^1^, Xingdong Chen^1^, Qiming Dai^1^, Wenxue Ding^1^, Yan Fang^1^, Jinwen Feng^1^, Lei Feng^1^, Shujing Fu^1^, Jian Gao^1^, Jieyu Ge^1^, Jiani Gu^1^, Qingxia Huang^1^, Wanwan Hou^1^, Liguo Jia^1^, Nan Jiang^1^, Wan Jiang^1^, Mengmeng Kong^1^, Xuan Lan^1^, Anxin Li^1^, Hao Li^1^, Jinxi Li^1^, Jing Li^1^, Ting Li^1^, Wenyu Li^1^, Yao Li^1^, Yingcen Li^1^, Huiqin Lin^1^, Quanzhong Liu^1^, Ting Liu^1^, Yangyang Liu^1^, Jingjing Luo^1^, Lan Luo^1^, Quanne Niu^1^, Yujie Pan^1^, Xiaolin Qian^1^, Xiaoqing Qiao^1^, Zhaoyu Qin^1^, Zhiwei Rong^1^, Bing Shen^1^, Shengli Shen^1^, Shiwei Shen^1^, Biru Shi^1^, Jing Sun^1^, Zhonghan Sun, Liangyan Tang^1^, Mengting Wan^1^, Chengyan Wang^1^, He Wang^1^, Jialin Wang^1^, Miaomiao Wang^1^, Qi Wang^1^, Qiying Wang^1^, Rui Wang^1^, Wenjing Zhang^1^, Yun Wang^1^, Yunzhi Wang^1^, Lingge Wu^1^, Xin Xia^1^, Hui Xie^1^, Binxin Xu^1^, Yongwu Xu^1^, Zhipeng Xu^1^, Liping Xue^1^, Jun Yan^1^, Cuiwei Yang^1^, Ying Yu^1^, Hui Yuan^1^, Chao Zhang^1^, Manfei Zhang^1^, Yining Zhao^1^, Yuanting Zheng^1^, Chen Zhu^1^, Lingli Zhu^1^

**Affiliations:**

^1^State Key Laboratory of Genetic Engineering, School of Life Sciences, Human Phenome Institute and Center for Evolutionary Biology, Fudan University, Shanghai 200433, China

## CONSORTIA

The members of the Human Phenome Project Scientific Consortium are Jun Chang, Wei Chen, Chen Ding, Li Jin, Tiemin Liu, Jingchun Luo, Qiang Luo, Yuan Qi, Feng Qian, Leming Shi, Deian Ta, Huiru Tang, Weishuo Tao, Mei Tian, Jiucun Wang, Sijia Wang, Shuhua Xu, Zhiqiang Yan, Zhong Yang, Guoqing Zhang, Menghan Zhang, Yan Zheng, and Lei Zhu.

The members of the Human Phenome Project Technological Consortium are Ran An, Wenyan Chai, Chen Chen, Longlong Chen, Qinsheng Chen, Weixin Chen, Xingdong Chen, Qiming Dai, Wenxue Ding, Yan Fang, Jinwen Feng, Lei Feng, Shujing Fu, Jian Gao, Jieyu Ge, Jiani Gu, Qingxia Huang, Wanwan Hou, Liguo Jia, Nan Jiang, Wan Jiang, Mengmeng Kong, Xuan Lan, Anxin Li, Hao Li, Jinxi Li, Jing Li, Ting Li, Wenyu Li, Yao Li, Yingcen Li, Huiqin Lin, Quanzhong Liu, Ting Liu, Yangyang Liu, Jingjing Luo, Lan Luo, Quanne Niu, Yujie Pan, Xiaolin Qian, Xiaoqing Qiao, Zhaoyu Qin, Zhiwei Rong, Bing Shen, Shengli Shen, Shiwei Shen, Biru Shi, Jing Sun, Zhonghan Sun, Liangyan Tang, Mengting Wan, Chengyan Wang, He Wang, Jialin Wang, Miaomiao Wang, Qi Wang, Qiying Wang, Rui Wang, Wenjing Zhang, Yun Wang, Yunzhi Wang, Lingge Wu, Xin Xia, Hui Xie, Binxin Xu, Yongwu Xu, Zhipeng Xu, Liping Xue, Jun Yan, Cuiwei Yang, Ying Yu, Hui Yuan, Chao Zhang, Manfei Zhang, Yining Zhao, Yuanting Zheng, Chen Zhu, and Lingli Zhu.

## Supporting information

Source Data for Figures 1-5 and Supplementary Figures S1-S11

## Data Availability

The multi-omics dataset of the ChiHOPE cohort (genomics, methylomics, miRNAomic, proteomic, targeted and untargeted metabolomic data) has been deposited at Genome Sequence Archive (GSA) as PRJCA026766 and at National Omics Data Encyclopedia (NODE, https://www.biosino.org/node) as OEP00002924. The multi-omics dataset of the NSPT cohort (miRNAomic and targeted metabolomic data) has been deposited at GSA as PRJCA033704 and at NODE as OEP00005910. Source data underlying the figures are provided with this article. The original analysis code was deposited at Zenodo (10.5281/zenodo.23991482). Any additional information required to reanalyze the data reported in this paper is available from the lead contact upon request.

https://doi.org/10.5281/zenodo.23991482

https://www.biosino.org/node

## Acknowledgments and funding

We gratefully acknowledge contributions from the Human Phenome Project Scientific Consortium (HPPSC) and the Human Phenome Project Technological Consortium (HPPTC) for their invaluable contributions to the design and execution of this project. We are grateful to CFFF (Computing for the Future at Fudan), the Human Phenome Data Center of Fudan University and the School of Public Health of Fudan University for computing support. Prof Huiru Tang at Fudan University is acknowledged for his valuable discussion. We thank the National Omics Data Encyclopedia (NODE, https://www.biosino.org/node/) by Bio-Med Big Data Center for help in data management support. We thank all the patients, physicians, and nurses who participated in this study. This study was supported by the National Natural Science Foundation of China (T2425013 to Y.T.Z., 32370701 to L.S., and 32470692 to Y.T.Z.), Shanghai Municipal Science and Technology Major Project (2023SHZDZX02 and 2017SHZDZX01 to L.J.), the National Key R&D Project of China (2023YFC3402501 to L.S.), the Strategic Priority Research Program of the Chinese Academy of Sciences (XDA12040104 to Y.T.Z.), State Key Laboratory of Genetic Engineering (SKLGE-2117 to L.S.), and the 111 Project (B13016 to L.S.).

## Author contributions

L.S., Y.T.Z., X.L. and A.V.P. conceived and oversaw the study. Y.T.Z. and L.S. coordinated multi-omics data generation and data analysis. D.S.P., Z.N., X.L., L.N.J., and W.J. coordinated sample collection during clinical trials. Z.C. conducted most of the data analysis and generated most of the figures. H.C., J.L., N.Z., Q.C., Y.M., X.Y., Y.L., Y.Y., J.T., X.C., Y.Z., H.W., J.W., J.C., Q.H., Q.W., S.W., Q.P., W.L., D.J.P., Y.W., T.X., A.V.P., and L.J. participated in data analysis, validation, or interpretation. Y.T.Z., Z.C., N.Z., Q.C., X.Y., S.S., Y.A, Z.Y., C.D., and B.J. generated the multi-omics datasets. Z.C., Y.T.Z., L.S. and A.V.P. wrote and revised the manuscript. All authors reviewed and approved the manuscript.

## Declaration of interests

D.S.P., Z.N., and X.L. are employees and stockholders of Shenzhen Chipscreen Biosciences Co. Ltd. The other authors declare no competing interests.

## Declaration of generative AI and AI-assisted technologies in the writing process

During the preparation of this work, the authors used Grammarly, Claude (Anthropic) and ChatGPT (OpenAI) to edit language and improve readability. After using these tools, the authors reviewed and edited the content as needed and take full responsibility for the content of the publication.

## Methods

### Study participants

#### T2D patients from the ChiHOPE cohort

The ChiHOPE cohort of 835 participants was constructed from patients with T2D recruited from two multi-center, randomized, double-blind phase 3 trials of chiglitazar (CMAP, placebo-controlled, ClinTrials.gov registration no. NCT02121717; CMAS, sitagliptin-controlled, ClinTrials.gov registration no. NCT02173457). The inclusion criteria were described in detail elsewhere^85,86^. Briefly, patients with recent-onset T2D were eligible for inclusion if they were aged 18-70 years with a BMI range of 18.5-35.0 kg/m^2^, and with insufficient glycemic control (HbA_1c_ ≥ 58.5 mmol/mol [7.5%] and ≤ 85.8 mmol/mol [10.0%]) despite a strict diet and exercise regimen^85,86^. The trial protocols excluded participants with T1D, refractory hypertension, triglyceride > 5.65 mmol/L, history of acute and chronic complications, and participants who have received oral or injectable hypoglycemic medication before recruitment, currently on fibrates, currently or have received any treatments that may change glucose metabolism^85,86^.

After two weeks of placebo run-in treatment, 535 patients from the CMAP trial were randomly assigned to receive chiglitazar 32 mg/day (*n* = 167), chiglitazar 48 mg/day (*n* = 166), or placebo (*n* = 202), while 739 patients from the CMAS trial were randomly assigned to receive chiglitazar 32 mg/day (*n* = 245), chiglitazar 48 mg/day (*n* = 246), or sitagliptin 100 mg/day (*n* = 248). Of the 1,274 patients participating in the two trials, 835 (65.5%) patients gave further consent for this companion multi-omics study, including 350 patients from the CMAP trial (chiglitazar 32 mg/day, *n* = 106; chiglitazar 48 mg/day, *n* = 106; placebo, *n* = 138) and 485 patients from the CMAS trial (chiglitazar 32 mg/day, *n* = 161; chiglitazar 48 mg/day, *n* = 161; sitagliptin, *n* = 163), which constitute the ChiHOPE cohort. Among the 835 T2D patients from the ChiHOPE cohort, 761 (91.1%) had clinical data at week 24, 35 (4.2%) had data at week 12 but not week 24, and 39 (4.7%) had baseline data only (**Table S1**, **Figure 1**).

Ethical approval was obtained from the Ethical Committees of the study centers. All procedures performed in the study involving human participants were in accordance with the ethical standards of the institutional and (or) national research committee, and with the Declaration of Helsinki and its later amendments or comparable ethical standards. All participants provided written informed consent.

#### Analysis populations from the ChiHOPE cohort

The full companion cohort comprised 835 participants. The development of molecular HOMA-IR models was restricted to 705 participants with eligible baseline molecular (targeted, untargeted metabolomics, and proteomics) and clinical data (sex, age, BMI, waist circumference, HOMA-IR, triglycerides, HDL cholesterol and ALT). Week 24 molecular HOMA-IR estimates were calculated for 593 participants with eligible molecular and clinical data at both baseline and week 24. Sensitivity analyses with additional adjustment for the first five ancestry principal components (PCs) were restricted to participants with complete PC data (week 0: *n* = 686, week 24: *n* = 577).

#### Participants from the NSPT cohort

The National Survey of Physical Traits (NSPT) cohort consists of 1,027 Han Chinese volunteers recruited from Zhengzhou, Henan, China. The external phenotype-association analysis used 333 participants with available age, sex, BMI, and relevant molecular and clinical data. The clinical characteristics of included participants are shown in **Table S1**. NSPT is a sub-project of the National Science & Technology Basic Research Project approved by the Ethics Committee of Human Genetic Resources of the School of Life Sciences, Fudan University, Shanghai (14117)^114^.

### Phenotypic outcomes

#### Insulin resistance (IR)

HOMA-IR is calculated using the following formula: fasting insulin (μU ml^−1^) × fasting glucose (mg dl^−1^) / 405^85,86^. We defined IR as HOMA-IR ≥ 4.31 for males and ≥ 4.51 for females, as set for the Chinese population^88^. HOMA-IR was available at baseline for 826 of 835 participants. Molecular scores were predicted on the log-transformed HOMA-IR value. We compared measured and molecular IR thresholds on the same scale, as described below.

#### BMI categories

Baseline BMI categories were defined according to the 2000 Asia-Pacific recommendations co-sponsored by the World Health Organization (WHO) Regional Office for the Western Pacific: normal weight (18.5 to < 23 kg/m^2^), overweight (23 to < 25 kg/m^2^) and obesity (≥ 25 kg/m^2^)^87^.

#### Metabolic syndrome (MetS) and non-glycemic components

Metabolic syndrome (MetS) was implemented using available measurements and the thresholds in the Chinese Diabetes Society 2024 guideline^115^. All participants had known T2D, so we considered the glycemic component present at every visit. Non-glycemic components were abdominal obesity (waist circumference ≥ 90 cm in men or ≥ 85 cm in women), elevated blood pressure (systolic pressure ≥ 130 mmHg or diastolic pressure ≥ 85 mmHg), elevated triglycerides (≥ 1.70 mmol/L) and low HDL cholesterol (< 1.04 mmol/L). MetS therefore required at least two of the four non-glycemic components. The non-glycemic component count ranged from zero to four, and required all four components to be observed. Missing components were not treated as absent, and MetS status was assigned from incomplete measurements only when the observed components were sufficient to determine it. Hypertension history, antihypertensive treatment, and lipid-lowering medication were unavailable, so the blood-pressure, triglyceride or HDL components were only based on measured values.

### Generation and quality control of longitudinal multi-omics datasets

Sample availability for the longitudinal multi-omics dataset is shown in **Fig. S1A-B**. For genomics and methylomics data, we collected whole blood samples from 834 T2D patients in the ChiHOPE cohort at baseline. For miRNAomic, proteomic, and metabolomic data, a total of 2,190 fasting plasma samples were collected at baseline, 12 weeks, and 24 weeks after treatment, from 756 T2D patients of the ChiHOPE cohort. Specifically, 1,990, 1,417, 2,148, and 2,063 plasma samples were used to generate miRNAomic, proteomic, targeted metabolomic, and untargeted metabolomic data, respectively.

#### Genomics

DNA was extracted from whole blood samples of 834 T2D patients of the ChiHOPE cohort at baseline (**Fig. S1A-B**). To ensure reliability, QC samples were included, comprising four types of Quartet DNA reference materials (D5, D6, F7, M8)^90^ and the RM8398 DNA reference material from the National Institute of Standards & Technology (NIST). DNA concentrations were quantified with a Qubit^®^ 3.0 Fluorometer (Thermo Fisher Scientific) and a NanoDrop One (Thermo Fisher Scientific), and DNA integrity was assessed by agarose gel electrophoresis.

DNA samples were placed on 96-well plates, with each plate containing at least four Quartet DNA reference materials and one RM8398 DNA reference material. DNA samples were genotyped with the Affymetrix Axiom Precision Medicine Research Arrays (Thermo Fisher Scientific) according to the manufacturer’s instructions. Raw data (CEL format) were generated by the Affymetrix GeneTitan workflow, and subsequently processed through the Best Practices Workflow embedded in Axiom Analysis Suite to call genotypes on 902,560 probes. All ChiHOPE samples (*n* = 834) met the QC threshold, and their predicted sex matched clinical records. After filtering duplicated and low detection quality SNPs, 874,438 SNPs were retained for downstream analysis.

Principal component analysis (PCA) of patient genomic data was performed using PLINK v1.90b6.27 (https://www.cog-genomics.org/plink/1.9/)^116^. Autosomal SNPs were filtered for genotype missingness (*--geno* 0.02), minor allele frequency (*--maf* 0.05), and Hardy–Weinberg equilibrium (*--hwe* 1e-6), followed by linkage disequilibrium (LD) pruning (*--indep-pairwise* 50 5 0.2). Individuals with heterozygosity rates more than three standard deviations above or below the sample mean were excluded (*n* = 12). Pairwise relatedness was estimated using LD-pruned SNPs (*--genome --min* 0.2), and the individual with the higher genotype missingness rate in each flagged pair was excluded (*n* = 8). Following these exclusions, 814 of the initial 834 individuals remained. The first five principal components were calculated using the LD-pruned SNPs (*--pca* 5), and were included as covariates in sensitivity analyses.

#### Methylomics

DNA was extracted from whole blood samples of 834 T2D patients in the ChiHOPE cohort at baseline (see the Genomics section **and Fig. S1A-B**). QC samples were included, comprising four types of Quartet DNA reference materials. Genome-wide DNA methylation was profiled using the Infinium MethylationEPIC BeadChips (Illumina) according to the manufacturer’s instructions. DNA samples were placed on 96-well plates, with each plate containing at least one Quartet DNA reference material. Genomic DNA was bisulfite-converted with the EZ DNA Methylation-GoldTM Kit (Zymo Research) and was hybridized to BeadChips. Single-base extension was performed using a Freedom EVO robot (Tecan). BeadChips were imaged using the iScan Microarray Scanner (Illumina) to generate raw data (IDAT format).

Raw data were processed with the R package minfi (v.1.48.0)^117^ without background correction^118^. Sex was predicted by the *getSex* function from minfi, which was concordant with the clinical records. Probes located near SNPs were identified and removed by the *dropLociWithSnps* function from minfi, and probes on sex chromosomes were also removed. The *detectionP* function from minfi was applied to test whether probe signals were significantly different from background noise. The *β* values with detection *P*-values greater than 0.05 were set as missing values, and 815,460 probes with no more than 5% missing values were retained. The missing *β* values were then imputed with the *impute.knn* function from the R package impute (v.1.78.0)^119^, based on the K nearest neighbors (KNN) algorithm (*k* = 5). The probe bias was adjusted with the beta mixture quantile (BMIQ) normalization method^120^ by the *champ.norm* function from the R package ChAMP (v.2.24.0)^121^. Singular value decomposition (SVD) was performed on the *β* value matrix using the *champ.SVD* function from ChAMP, revealing a significant batch effect on PC2 (explaining 18.5% of the variance). Batch effects were corrected using the ComBat function^122^ from the R package sva (v.3.52.0)^123^ in M values (logit-transformed *β* values), and then the M values were transformed back to *β* values.

The cell composition proportions of whole blood samples of 834 T2D patients from the ChiHOPE cohort were estimated from the methylomic data using the *epidish* function (*ref.m* = centDHSbloodDMC.m, *method* = RPC) from the R package EpiDISH (v2.10.0)^124^. Other parameters were set to their default values.

Eleven established DNA methylation-derived measures were calculated from the processed baseline blood methylation *β*-value matrix using frozen published models: Horvath pan-tissue age^125^, Hannum age^126^, Horvath skin-and-blood age^127^, PC-Horvath age^128^, AltumAge^129^, PhenoAge^130^, GrimAge versions 1 and 2^103,131^, DunedinPoAm^132^, DunedinPACE^99^ and DNA methylation-estimated telomere length (DNAmTL)^133^. Clock coefficients, PC loadings, scaling references and neural-network weights were obtained from an archived Biolearn reference snapshot (commit 0d714f5a0c0ad5743b6315f3edab1f86112d394f) and applied with a reproducible Python implementation. Linear clocks used the published weighted sums and inverse age transformations where required. PC-Horvath used its published centering vector, principal-component loadings and prediction coefficients. AltumAge used the published input scaling and frozen network weights. GrimAge models combined their methylation surrogates with chronological age and sex according to the published algorithms. For unavailable model CpGs, the implementation used reference methylation values from the Biolearn sesame_450k_median reference; sporadic missing values at CpGs observed in at least 80% of samples were filled with the observed CpG mean. DunedinPACE instead used its own published gold-standard reference means and target-distribution normalization over the required 20,000-CpG background before applying the 173-CpG prediction model. DNAmTL used the corrected positive intercept of 7.924780053.

#### MiRNAomics

Total RNA was extracted from 1,990 fasting plasma samples of T2D patients in the ChiHOPE cohort (week 0, *n* = 679; week 12, *n* = 667; and week 24, *n* = 644) and 993 fasting plasma samples from individuals in the NSPT cohort (**Fig. S1A-C**). QC samples were included to ensure sequencing reliability, comprising plasma samples from a healthy individual (P10, *n* = 31) and mixed plasma samples from ChiHOPE patients (PM, *n* = 34). Plasma samples were aliquoted, and total RNA, including extracellular RNA (exRNA), was extracted using the miRNeasy Serum/Plasma Advanced Kit (QIAGEN). The exRNA library was prepared using the NEXTflex^®^ Small RNA-seq Kit v3 (PerkinElmer). RNA quantity and quality were assessed using a Qubit Flex fluorometer (Invitrogen) and an Agilent Bioanalyzer (Agilent Technologies) before library construction and sequencing. After final quantification using a Qubit Flex fluorometer (Invitrogen) and size distribution confirmation with the BiOptic Qsep100, all RNA and exRNA libraries underwent paired-end (150-bp reads) sequencing on the Illumina NovaSeq6000 platform (Illumina), with an average depth of 15.7M raw reads per library to generate the raw sequencing files (FASTQ format).

Raw data were processed with the ExceRpt small RNA sequencing data analysis pipeline (docker v4.6.3)^134^ for sequence alignment and counting. Default parameters were applied except for the following modifications. First, we used TGGAATTCTCGGGTGCCAAGG as the 3’ adapter sequence, overriding the adapter sequences the pipeline guessed. Second, we set the random barcode length to 4. Third, the priority of the reference libraries during read assignment was set to miRNA > piRNA > tRNA > GENCODE > circRNA^135^. The remaining reads were then aligned to the human reference genome (Genome Reference Consortium GRCh38, UCSC hg38) and miRBase version 21 (http://www.mirbase.org/), and the numbers of aligned reads for each miRNA were counted. Libraries with low sequencing depths (< 1M raw reads), mapping ratios lower than 50%, and low transcript-genome ratios were removed. A total of 396 highly detected miRNAs with at least 10 read counts in at least 50% of all samples were retained. We normalized read counts using the counts per million (CPM) method and then log-transformed them.

#### Proteomics

Peptides were extracted from 1,417 fasting plasma samples of T2D patients in the ChiHOPE cohort (week 0, *n* = 736; and week 24, *n* = 681) (**Fig. S1A-B, *F*-G**). QC samples were included, comprising plasma samples from two healthy individuals (P10, *n* = 16; and P11, *n* = 16) and the PM samples (*n* = 31). We diluted and alkylated plasma samples at 95°C for 10 min before protein extraction. We digested the depleted plasma samples with trypsin at an enzyme-to-protein mass ratio of 1:25 overnight at 37°C, then extracted and dried the peptides (SpeedVac, Eppendorf).

The LC-MS/MS instrumentation for measurement consisted of an EASY-nLC 1200 ultra-high-pressure system (Thermo Fisher Scientific). It was coupled to a Q Exactive HF-X (Thermo Fisher Scientific) via a nano-electrospray ion source. For each measurement, 2 μL of the purified peptides dissolved in 100 μL of loading buffer A (0.1% formic acid in water) was loaded onto a 75 μm I.D. × 10 cm C18 column (C18, 1.9 μm, 120 Å, Dr. Maisch GmbH) at a maximum pressure of 350 bar with 4 μL of solvent A (0.1% formic acid in water). Peptides were eluted with a linear gradient of 15–30% Mobile Phase B (acetonitrile and 0.1% formic acid) at 600 nL/min. The MS data were acquired in a data-independent acquisition (DIA) method, in which an MS1 scan was acquired from 300–1400 m/z at 30k resolution (gain control target [AGC] of 4 × 10^5^ or injection time [IT] of 50 ms). Then, we acquired 30 DIA segments at 15k resolution with an AGC of 1 × 10^6^ or an IT of 22 ms for maximal injection time, allowing ion injection for all available parallelizable time. We set higher-energy collisional dissociation (HCD) fragmentation to a normalized collision energy of 27%. The spectra were recorded in profile mode. The default charge state for the MS2 was set to 3.

All data were processed using Firmiana^136^. The DIA data were searched against the UniProt human protein database (updated on 2019.12.17, 20,406 entries) using FragPipe (v12.1) with MSFragger (2.2)^137^. We set mass tolerances of 20 ppm for precursor ions and 50 mmu for product ions. Up to two missed cleavages were allowed. The search engine set cysteine carbamidomethylation as a fixed modification and N-acetylation and methionine oxidation as variable modifications. Precursor ion charge states were limited to +2, +3, and +4. The data were also searched against a decoy database so that protein identifications were accepted at a false discovery rate (FDR) of 1%. The results of DIA data were combined into spectra libraries using SpectraST software. A total of 327 libraries were used as reference spectra libraries.

DIA data were analyzed using DIA-NN (v1.7.0)^138^. The default settings were used for DIA-NN (Precursor FDR: 5%, Log lev: 1, Mass accuracy: 20 ppm, MS1 accuracy: 10 ppm, Scan window: 30, Implicit protein group: genes, Quantification strategy: robust LC [high accuracy]). Quantification of identified peptides was calculated as the average of chromatographic fragment ion peak areas across all reference spectra libraries. Label-free protein quantifications were calculated using a label-free, intensity-based absolute quantification (iBAQ) approach^139^. We calculated peak areas as fractions of the corresponding proteins. The fraction of total (FOT) was used to represent the normalized abundance of a particular protein across samples. FOT was defined as a protein’s iBAQ divided by the total iBAQ of all identified proteins within a sample. We multiplied the FOT values by 10^5^ for ease of presentation. The peptide-to-protein inference algorithm xTop was used to combine intensities from unique peptides of a given protein across all samples at hand to infer the intensities of that protein, which outperformed other common protein inference algorithms such as TopPep3 and iBAQ in accuracy^140^.

A total of 837 proteins with less than 30% missing values across all samples were included. Protein intensities were log-transformed and then quantile-normalized. Batch effects were observed, with protein quantities distributed across loading-order groups identified as MS signal drifts. LOESS fitting in each separate group was conducted to correct each protein quantity^141^. We applied median centering, subtracting each peptide’s median quantity across samples in each group from its quantity per sample, to remove residual bias within discrete groups.

#### Untargeted metabolomics

A total of 2,063 fasting plasma samples of T2D patients in the ChiHOPE cohort (week 0, *n* = 713; week 12, *n* = 701; and week 24, *n* = 649) were used for untargeted metabolomics (**Fig. S1A-B, E**). The PM samples (*n* = 182) were included to ensure quantification reliability.

We mixed each plasma sample (180 μL) with 360 μL of phosphate buffer (45 mM, pH 7.43) in a 5 mm NMR tube and used it directly for NMR analysis. All NMR spectra were acquired at 298 K on a Bruker AVANCE III 600 MHz NMR spectrometer (600.13 MHz for ^1^H frequency) equipped with a quaternary cryogenic inverse probe (Bruker Biospin, Germany). For each plasma sample, three spectra were collected, including one with the standard NOESYPR1D sequence (RD−90°−*t*1−90°−tm−90°−acq), a *t*2-edited spectrum with a standard Carr−Purcell−Meibom−Gill (CPMG) sequence (RD−90°−(τ−180°−τ)*n*−acq) with τ of 350 μs and *n* of 100, and a diffusion-edited spectrum (with the sequence, RD−90°−G1−τ−180°−G2−τ−90°−Δ−90°−G3−τ−180°−G4−τ−90°−te−90°−acq) with Δ of 200 ms, τ of 150 μs, and te of 5 ms. While NOESYPR1D spectra contained all detectable organic metabolite signals, CPMG spectra contained mainly signals from small metabolites or fast-moving moieties. In contrast, the diffusion-edited spectra contained signals from lipid moieties in lipoproteins and acetyl-glycoproteins. The 90° pulse length was adjusted to about 10 μs for each sample, and 64 transients were collected into 32,000 data points over a spectral width of 20 ppm. For resonance assignment, we acquired a series of two-dimensional (2D) NMR spectra for selected samples and processed them as previously reported. These included ^1^H−^1^H correlation spectroscopy (COSY), ^1^H−^1^H total correlation spectroscopy (TOCSY), J-resolved spectroscopy (JRES), ^1^H−^13^C heteronuclear single quantum correlation (HSQC), and ^1^H−^13^C heteronuclear multiple bond correlation (HMBC) 2D NMR spectra. All the NMR spectra were processed using the software package TOPSPIN (V3.6.0, Bruker Biospin, Germany). For ^1^H NMR spectra, we applied an exponential window function with a line-broadening factor of 1 Hz and zero-filled to 128 k before Fourier transformation. Each spectrum was phase- and baseline-corrected manually and referenced to the anomeric proton signal of α-glucose (δ 5.23). We then divided the spectral regions of δ 0.5−9 into bins 0.002 ppm (1.2 Hz) wide using the AMIX software package (V3.9.15, Bruker Biospin). The integrals of the bins were normalized to the volume. The relative content of metabolites was calculated by the characteristic and least-overlapping NMR signals.

Untargeted metabolomic datasets included 30 small molecular and 10 large molecular metabolites (33 unique metabolites). Ratios between unsaturated fatty acids, polyunsaturated fatty acids, monounsaturated fatty acids and saturated fatty acids were calculated. Metabolomic data (original values, excluding ratios) were log-transformed.

#### Targeted metabolomics

A total of 2,148 fasting plasma samples of T2D patients in the ChiHOPE cohort (week 0, *n* = 737; week 12, *n* = 729; and week 24, *n* = 682) and 339 samples from individuals in the NSPT cohort were used for targeted metabolomics (**Fig. S1A-B, D**). QC samples, comprising P10 (*n* = 90), P11 (*n* = 45), and PM samples (*n* = 45), were included to ensure reliable quantification.

The metabolomics analysis was performed using the Q300 Kit (Metabo-Profile, Shanghai, China). Plasma samples were prepared following the manufacturer’s instructions. Samples were thawed in an ice bath to diminish sample degradation, and 25 μL of plasma was added to each well of a 96-well plate. The plate was then transferred to the Biomek 4000 workstation (Biomek 4000, Beckman Coulter, Inc., Brea, California, USA), where the instrument automatically added 100 μL of ice-cold methanol with partial internal standards to each sample and vigorously vortexed for 5 min. The plate was centrifuged at 4,000 × g for 30 min (Allegra X-15R, Beckman Coulter, Inc., Indianapolis, IN, USA) and then returned to the workstation. Next, 30 μL of supernatant was transferred to a clean 96-well plate, and 20 μL of freshly prepared derivative reagents was added to each well. The plate was sealed, and the derivatization was carried out at 30°C for 60 min. Following derivatization, 350 μL of ice-cold 50% methanol solution was added to dilute the samples. The plate was then stored at −20°C for 20 min and centrifuged at 4,000 × g for 30 min at 4 °C. A volume of 135 μL of supernatant was transferred to a new 96-well plate, with 15 μL of internal standards added to each well. Serial dilutions of derivatized stock standards were added to the left wells. Finally, the plate was sealed for LC-MS analysis. Sample acquisition was randomized to avoid bias.

Targeted metabolic profiling was conducted at Metabo-Profile (Shanghai, China) using an ultra-performance liquid chromatography coupled to tandem mass spectrometry (UPLC-MS/MS) system (ACQUITY UPLC-Xevo TQ-S, Waters Corp., Milford, MA, USA). The optimized instrument settings are as follows: For UPLC, column: ACQUITY UPLC BEH C18 1.7 × 10^−6^ m VanGuard precolumn (2.1 × 5 mm) and ACQUITY UPLC BEH C18 1.7 × 10^−6^ m analytical column (2.1 × 100 mm), column temp.: 40 °C, sample manager temp.: 10 °C, mobile phases: A = water with 0.1% formic acid; and B = acetonitrile / IPA (90:10), gradient conditions: 0-1 min (5% B), 1-12 min (5-80% B), 12-15 min (80-95% B), 15-16 min (95-100% B), 16-18 min (100% B), 18-18.1 min (100-5% B), 18.1-20 min (5% B), flow rate: 0.40 mL min^−1^, and injection volume: 5.0 µL. For MS, capillary: 1.5 (ESI+), 2.0 (ESI-) Kv, source temp.: 150 °C, desolvation temp.: 550 °C, and desolvation gas flow: 1000 L h^−1^.

The raw data generated by UPLC-MS/MS were processed using the QuanMET software v2.0 (Metabo-Profile, Shanghai, China) to perform peak integration, calibration, and quantitation for each metabolite. A total of 203 metabolites with less than 30% missing values across all samples were retained. Missing values were imputed with half of the minimum value for each metabolite across all samples, and the concentrations were log-transformed.

### Development of molecular HOMA-IR estimates

We developed molecular HOMA-IR models using baseline data from 705 ChiHOPE participants with metabolomic, proteomic, and clinical measurements. A total of 217 metabolites (202 targeted and 15 untargeted) and 837 proteins were included in the model development. Duplicated metabolites from untargeted metabolomic data and D-glucose were excluded. The prediction target was log-transformed HOMA-IR. Separate Gaussian elastic-net regression models^142,143^ were fitted to metabolomic and proteomic inputs to obtain met-HOMA-IR and prot-HOMA-IR, respectively. These two predictions were combined by ordinary least squares (OLS) regression to obtain mol-HOMA-IR.

#### Repeated nested cross-validation and molecular integration

Model development used 20 repetitions of participant-level ten-fold outer cross-validation, with identical outer assignments for all models. Fold construction balanced clinical trial, measured IR status, sex and BMI category. Age means and quartiles were additionally balanced across outer folds by exchanging participants within the same joint strata. The 705 participants retained their assignments from the predefined fold plan, and every model was refitted using only these participants. Each outer iteration held out about one-tenth of participants for evaluation and used the remainder for model development. We selected hyperparameters using five-fold cross-validation within each outer training set.

After the upstream assay processing described above, all model-specific imputation and standardization parameters were estimated from the relevant training fold. Features with fewer than two finite training observations or a training standard deviation of 10^−10^ or less were omitted. Missing values were imputed using the training median, and features were centered and scaled using the training mean and sample standard deviation. We also centered and scaled the target during fitting; predictions were then returned to log HOMA-IR units. We repeated these steps within each inner training fold and, after tuning, on the full outer training set. The corresponding parameters were then applied without re-estimation to the held-out data.

The elastic-net mixing parameter *α* was evaluated at 0.1, 0.3, 0.5, 0.7, 0.9 and 1.0, and the penalty parameter *λ* was evaluated at 60 logarithmically spaced values from 10 to 0.001. Models included an intercept. Predictive loss was the mean squared error (MSE) on the original log HOMA-IR scale, averaged across the five inner validation folds. The *α* value attaining the minimum MSE across the complete grid was selected. At this *α*, the primary model used the largest *λ* whose MSE was within one standard error of the minimum, where the standard error was the standard deviation of the five fold-specific MSEs divided by √5. Minimum-MSE models were also retained as secondary outputs.

For molecular integration, we performed an additional five-fold cross-fitting procedure entirely within each outer training set. In each cross-fitting split, both single-omics models were retuned using five-fold inner cross-validation, fitted to the cross-fitting training subset and applied to its held-out subset. The resulting training-set cross-fitted predictions were entered into an OLS model, log HOMA-IR = *b*_0_ + *b*_M_ × met-HOMA-IR + *b*_P_ × prot-HOMA-IR. The fitted intercept and weights were then applied to the two outer-test single-omics predictions. We treated a base score with negligible variation as constant to avoid unstable integration weights. Outer-test participants were excluded from model-stage preprocessing estimation, hyperparameter selection, base-model fitting and estimation of the integration weights.

Each participant received one outer-held-out prediction for each score in each repetition. The 20 predictions were averaged on the log scale to obtain a single baseline estimate per participant. For longitudinal evaluation, we applied each frozen baseline outer-fold model, including its preprocessing parameters and integration weights, to week-24 molecular measurements from the same held-out participants. No week-24 data were used for model training or recalibration. The common week-24 evaluation population comprised 593 participants, and the 20 week-24 predictions were averaged in the same manner.

#### Prediction performance and clinical benchmarks

For each cross-validation repetition, predictions from all ten outer folds were pooled, and out-of-sample *R*^2^ was calculated as 1 − Σ(y − ŷ)^2^/Σ(y − ȳ)^2^, where y was measured log HOMA-IR. *R*^2^ was summarized across the 20 repetitions. Clinical benchmarks were fitted and evaluated on the same participants and outer folds. The triglyceride-to-HDL cholesterol ratio and triglyceride-glucose (TyG) index were each used as a single predictor in OLS models of log HOMA-IR. The ratio was log-transformed, and TyG was defined as ln[triglycerides (mg/dl) × fasting plasma glucose (mg/dl)/2]. The clinical model included BMI, waist circumference, log triglycerides, log HDL cholesterol, and log alanine aminotransferase. The combined model additionally included mol-HOMA-IR. Its training-set molecular covariate was generated by cross-fitting the entire base-model and integration procedure within each outer training set.

Model comparisons used approximate two-sided Nadeau–Bengio corrected paired *t*-tests^144^ on the 200 paired outer-fold MSE differences. The corrected standard error was √[(1/200 + ρ) × Var(d)], where d denoted fold-specific MSE differences and ρ was the mean baseline test-to-training sample-size ratio; tests used 199 degrees of freedom. Frozen week-24 comparisons used the same baseline training-overlap factor. BH correction^145^ was applied separately by visit across three comparisons among the molecular estimates, ten comparisons among the five models in the clinical-benchmark comparison, and three full-versus-restricted comparisons. Pearson correlations between measured HOMA-IR and participant-mean molecular predictions used two-sided tests with BH correction across the three molecular estimates at each visit.

An exploratory analysis used 654 participants with baseline metabolomic, proteomic and miRNAomic data; participants missing the entire miRNA block were excluded. Single-omics models were refitted in this matched population using 20 repetitions of ten-fold outer cross-validation and five-fold inner tuning, retaining the existing participant fold assignments. Training-set cross-fitted predictions were combined by OLS to produce either a three-omics model or a matched metabolomic–proteomic reference. Direct prediction models for log HOMA-IR used the one-standard-error tuning rule. Out-of-sample *R*^2^ was compared at baseline (*n* = 654) and week 24 (*n* = 547), applying the frozen baseline models to follow-up measurements. This matched three-omics comparison was separate from the primary analysis of 705 participants at baseline and 593 at week 24. Circulating miRNAs were not included in the final molecular HOMA-IR estimates, because no improvement in model performance was seen after adding miRNAs to the integrated model.

#### Molecular coefficients, selection stability and single-analyte explained variance

Single-omics coefficients were expressed as the change in predicted log HOMA-IR per training-fold standard deviation of the molecular feature. Effective coefficients in mol-HOMA-IR were obtained by multiplying each single-omics coefficient by the corresponding outer-fold integration weight. Non-selected coefficients were assigned zero before calculating summaries across the 200 outer fits. We assessed selection stability both across all fits and within each of the 20 repetitions. For the broader single-analyte analysis, molecules were required to have a nonzero coefficient in at least one of the ten outer-fold models in every cross-validation repetition, evaluated separately for the metabolomic and proteomic models. This criterion identified 83 metabolites and 65 proteins. For each analyte, we regressed log HOMA-IR on that analyte using OLS with an intercept and no additional covariates, using the same 20 repetitions of ten-fold outer cross-validation. Missing values were imputed from the training-fold median. Fold-specific models were applied to held-out participants, and the pooled out-of-sample *R*^2^ was calculated for each repetition.

#### Restricted models and external phenotype associations in NSPT

For application to NSPT, restricted models used a fixed shared panel of 202 targeted metabolites and 754 proteins available across the cohorts. The predefined panel retained proteins with less than 70% missingness in both the development reference population (*n* = 706) and external reference population (*n* = 333). Restricted models were retuned and refitted on the same 705 development participants and identical cross-validation assignments as the full models. Their performance was compared using the same baseline and week-24 evaluation populations.

For deployment to new participants, the restricted single-omics models were retuned and fitted using all 705 baseline participants. We estimated final integration weights by OLS using pooled, repeated baseline out-of-fold single-omics predictions, with equal total weight per participant. These deployment models were used only for external prediction, not for the reported baseline or week-24 cross-validation performance. Before prediction in the 333 eligible NSPT participants, each active molecular feature underwent an unsupervised cohort-level location and scale transformation, X* = μtraining + σtraining × (Xexternal − μexternal)/σexternal. Training statistics were calculated using the 705-person development cohort, and external statistics used observed values in the full eligible molecular reference cohort before phenotype-specific exclusions. We then applied frozen development preprocessing and model coefficients, including training-median imputation of missing values. This procedure provided exploratory cohort adaptation rather than assay calibration.

Because NSPT did not measure HOMA-IR, external evaluation examined associations with clinical phenotypes rather than predictive accuracy for HOMA-IR. OLS models related each clinical phenotype to each molecular estimate, adjusting for age and sex. We standardized estimates using the score standard deviation across all 333 participants and the phenotype standard deviation within each phenotype’s eligible sample. We log-transformed fasting plasma glucose, triglycerides, HDL cholesterol, the triglyceride-to-HDL cholesterol ratio, alanine aminotransferase, aspartate aminotransferase, and the aspartate-to-alanine aminotransferase ratio. BMI, waist and hip circumference, waist-to-hip ratio, body-fat and visceral-fat measures, and systolic and diastolic blood pressure were analyzed on their original scales before standardization. The original phenotype-specific quality-control and outlier masks were retained. Inference used HC3 heteroscedasticity-robust standard errors and two-sided *t*-tests; BH adjustment covered 45 tests (15 phenotypes × three estimates).

#### Sensitivity models including BMI as an unpenalized covariate

To assess sensitivity to explicit adiposity information in model development, we added BMI to each single-omics model with a penalty factor of zero, whereas molecular features retained a penalty factor of one. Models used the same 705 participants, outer assignments, training-fold preprocessing and hyperparameter grid as the primary models, and were retuned within each training set. We fitted the integrated model using OLS with cross-fitted BMI-augmented single-omics predictions and BMI as predictors. BMI was included at the visit being scored; therefore, week-24 predictions used contemporaneous week-24 BMI. A BMI-only OLS model was evaluated on the same folds. Participant scores and calibrated gaps were obtained using the same averaging and training-set calibration procedures as above. We summarized differences in mean repeated-cross-validation *R*^2^ with 95% percentile confidence intervals from 2,000 paired participant bootstrap samples, retaining all 20 predictions from each sampled participant. These intervals condition on the already fitted models and do not incorporate uncertainty from refitting the complete training procedure.

#### Molecular-feature deletion analyses

We re-evaluated feature eligibility after each refitting round, requiring selection in at least eight of ten outer folds in each repetition, with selection in all ten folds as a sensitivity criterion. Among eligible features, we removed the feature with the largest absolute mean signed coefficient across the 200 fitted models, including zeros. We retuned and refit the affected base models on the original participant splits, and we re-estimated integration weights from training-only cross-fitted predictions for the molecular model. We continued this procedure for up to 12 deletions, stopping earlier if no eligible feature remained. Performance was summarized as pooled out-of-fold *R*^2^ within each repetition. Because the deletion sequence was selected using coefficients summarized across all outer models, these curves were treated as exploratory model-perturbation analyses rather than unbiased validation of a newly selected reduced panel.

### Statistical analysis

#### Molecular HOMA-IR gaps

Gaps were defined as calibrated residual deviations of the molecular estimate from the value expected for measured log HOMA-IR. For each single-omics score and outer split, the training-only cross-fitted predictions described above were regressed on measured log HOMA-IR: score = *a* + *b* × log HOMA-IR. For a held-out participant, the gap was calculated as predicted log HOMA-IR − *a* − *b* × log HOMA-IR. For mol-HOMA-IR, the cross-fitted single-omics predictions were combined using the integration weights fitted in that outer training set, and this fixed-weight training score was regressed on measured log HOMA-IR. The single-omics predictions were cross-fitted, whereas the integration weights used for gap calibration were estimated from the same outer-training cross-fit data. Calibration coefficients were fitted without outer-test participants. At week 24, the frozen baseline calibration coefficients were applied to the week-24 molecular estimate and contemporaneous measured log HOMA-IR. Gaps were averaged across the 20 repetitions for each participant and visit. This definition differs from simply subtracting measured from predicted log HOMA-IR.

#### Associations of molecular HOMA-IR estimates and gaps with clinical and epigenetic measures

We evaluated contemporaneous associations separately at baseline and week 24 using available observations from the 705-participant baseline and 593-participant week-24 cohorts, respectively. The clinical analysis included 18 measures: BMI, waist circumference, 2-hour postprandial glucose, HbA1c, triglycerides, HDL cholesterol, the triglyceride-to-HDL cholesterol ratio, total cholesterol, free fatty acids, alanine aminotransferase, aspartate aminotransferase, the aspartate-to-alanine aminotransferase ratio, urinary albumin-to-creatinine ratio, estimated glomerular filtration rate, high-sensitivity C-reactive protein, white blood cell count, systolic blood pressure and diastolic blood pressure. Clinical outcomes were natural-log transformed and standardized using the mean and sample standard deviation within the eligible analysis sample. Measured log HOMA-IR and each of the three molecular HOMA-IR estimates were standardized and analyzed as separate predictors in OLS models adjusted for sex, age and clinical trial. Epigenetic analyses used baseline measurements of Horvath v1, Hannum, Horvath v2, PC Horvath1, AltumAge, PhenoAge, GrimAge v1, GrimAge v2, DunedinPoAm38, DunedinPACE and DNA methylation-based telomere length (DNAmTL). These 11 outcomes were analyzed on their original scales, without log transformation or prior residualization on chronological age. Models included the covariates above and five estimated blood-cell fractions: B cells, natural killer cells, CD4 T cells, CD8 T cells and monocytes.

For gap analyses, each molecular gap was entered separately in place of the molecular estimate, and measured log HOMA-IR was included as an additional covariate. The clinical model was z[ln(clinical measure)] ∼ z(molecular HOMA-IR gap) + measured log HOMA-IR + sex + age + trial. Epigenetic models additionally included the five cell fractions. Gaps were standardized directly without a further log transformation. Primary inference used conventional OLS two-sided *t*-tests and 95% confidence intervals. HC3 heteroscedasticity-robust *t*-tests were supplied as sensitivity analyses. Multiple testing was controlled using the BH method separately by visit and outcome domain. Score analyses included 72 clinical tests per visit (18 measures × four IR estimates) and 44 baseline epigenetic tests (11 measures × four estimates). Gap analyses included 54 clinical tests per visit and 33 baseline epigenetic tests. OLS and HC3 results were adjusted separately.

#### Clinical and epigenetic measures across measured and molecular IR categories

For each molecular estimate and visit, measured and molecular IR were defined using the same sex-specific thresholds, applied on the log scale to the mean molecular prediction across repetitions. Their joint classification defined four categories: IR−/mol-IR−, IR−/mol-IR+, IR+/mol-IR− and IR+/mol-IR+. Clinical and epigenetic outcomes were analyzed using OLS with the four-category factor, sex, age and trial; epigenetic models additionally included the five blood-cell fractions. Clinical outcomes were natural-log transformed before standardization, whereas epigenetic outcomes were standardized without log transformation. Analyses used outcome-specific available observations without additional imputation. Adjusted means were standardized to the eligible sample’s covariate distribution; exponentiating adjusted clinical log means and confidence limits yielded geometric means and 95% confidence intervals. Overall and pairwise differences were assessed using three-degree-of-freedom *F*-tests and two-sided *t*-tests. BH correction covered 54 clinical overall tests and 324 clinical pairwise comparisons across 18 outcomes and three molecular classifications, and 33 epigenetic overall tests and 198 pairwise comparisons across 11 outcomes and three classifications, separately by visit and domain.

MetS and each binary non-glycemic component were modeled using logistic regression with the same four-category factor, sex, age and trial. Overall differences used three-degree-of-freedom likelihood-ratio tests and pairwise differences used two-sided Wald tests. Component counts were modeled using OLS with HC3 robust *F*- and *t*-tests. BH correction covered 18 overall tests and 108 pairwise comparisons across six outcomes (MetS, the component count and four binary components) and three molecular classifications, separately by visit. Stacked bars showed observed distributions; their annotated tests came from the adjusted models.

Adiposity sensitivity analyses additionally adjusted gap and four-category models for contemporaneous BMI and waist circumference. These two variables were omitted as clinical outcomes. Gap analyses then used 48 clinical and 33 epigenetic tests; four-category analyses used 48 clinical overall tests and 288 pairwise comparisons, with the epigenetic testing families unchanged. MetS retained its original definition including waist circumference, but the separate non-glycemic count excluded waist circumference and ranged from zero to three. The corresponding MetS-related family comprised five outcomes and three molecular classifications (15 overall tests and 90 pairwise comparisons). The same downstream adiposity adjustment was applied to gaps and classifications derived from the BMI-unpenalized models.

#### Selection-bias and inverse-probability-weighting analyses

Baseline characteristics were compared between included and excluded participants for the nested 835-, 705- and 593-person populations. Continuous and binary differences were assessed using two-sided Mann–Whitney U and Fisher’s exact tests, respectively, and standardized mean differences quantified imbalance. Exploratory *P* values were BH-adjusted within-trial and selection comparisons. Inverse probability weighting (IPW) examined selection of the 705 or 593 participants from the full cohort and selection of the 593 from the 705-person cohort.

Selection probabilities were estimated by bias-reduced logistic regression^146^. Core models contained the six trial-by-treatment strata, sex, age, BMI, waist circumference, systolic and diastolic blood pressure. Models within the 705-person cohort additionally included baseline log HOMA-IR, log triglycerides, log HDL cholesterol, log ALT, log fasting glucose, HbA1c and mol-HOMA-IR. HbA1c was complete in the 705-person cohort. Stabilized inverse selection weights were calculated as the marginal selection probability divided by the fitted individual probability and normalized to a mean of one. Untruncated weights and weights truncated at the first and 99th percentiles were evaluated. Weight distributions, effective sample sizes and covariate balance were inspected.

Weighted and unweighted week-24 ANCOVA estimates were compared for measured and molecular HOMA-IR. Outcome models adjusted for the corresponding baseline score, sex and age, without ancestry-PC adjustment. Score-specific IR-reversal risks and between-arm risk differences among participants positive at baseline were calculated directly from weighted or unweighted observations, without additional regression adjustment. Conditional HC3 standard errors were calculated for the continuous outcome models. For both types of outcome, uncertainty was assessed using 1,000 participant bootstrap resamples within trial-by-treatment strata. Each bootstrap refitted both the selection and outcome models while retaining the frozen molecular predictions. We obtained percentile confidence intervals and bootstrap-standard-error-based two-sided *P* values, with BH adjustment within the corresponding population, selection model, weighting variant and outcome family. This procedure assessed sensitivity to measured selection predictors but did not eliminate potential bias from unmeasured determinants of missingness.

#### Baseline treatment-group balance and week-24 molecular response

Baseline mol-HOMA-IR estimates were compared among treatment arms within each trial using Welch one-way ANOVA, with two-sided Welch *t*-tests for pairwise comparisons. The four-category distribution and marginal mol-IR status were compared using Pearson chi-square tests; four-category pairwise comparisons used the same test. Pairwise *P* values were Holm-adjusted across the three arm pairs within each trial and outcome. These exploratory baseline comparisons were unadjusted for covariates.

Treatment effects on measured and molecular HOMA-IR were evaluated among 593 paired participants (CMAP, *n* = 219; CMAS, *n* = 374). Within each trial, ANCOVA modeled the week-24 score using treatment arm, the corresponding baseline score, sex and age. Measured HOMA-IR was natural-log transformed; molecular estimates were already on the log-HOMA-IR scale. Adjusted mean changes were calculated as standardized week-24 predicted means minus the trial-specific mean baseline score. Overall and pairwise treatment differences used partial *F*-tests and two-sided *t*-tests.

Week-24 four-category outcomes were analyzed using mean-bias-reduced multinomial logistic regression^147^ with treatment arm, baseline four-category status, sex and age as predictors. Adjusted probabilities were obtained by averaging counterfactual predictions over the trial-specific analysis sample. Overall and pairwise treatment differences used joint Wald tests with six and three degrees of freedom, respectively. BH correction covered seven overall tests and 21 pairwise comparisons within each trial across four continuous HOMA-IR measures and three molecular classifications. These probability analyses included all paired participants irrespective of baseline status, and described week-24 states rather than reversal restricted to participants positive at baseline. Treatment-effect models did not include BMI change.

#### Decomposition of treatment-associated molecular changes

For participant i and feature j, we calculated the feature contribution to the mol-HOMA-IR change within each held-out baseline model as its effective frozen coefficient multiplied by the change in the corresponding preprocessed molecular measurement. Contributions were averaged across the 20 repetitions. The effective coefficient incorporated the single-omics coefficient and its integration weight. Because preprocessing and coefficients were frozen, the sum of the feature contributions reconstructed the individual predicted score change. The candidate set comprised 83 metabolites and 65 proteins selected in at least one of ten outer folds in every repetition. Within each trial, each feature contribution was regressed on treatment arm, baseline mol-HOMA-IR, sex and age. For each dose-control contrast, the percentage contribution to a lower score was 100 × max(−δj, 0)/Σk max(−δk, 0), where δj was the adjusted treatment contrast for feature j. The denominator included all modeled molecular features, including those not displayed. Features with negative contributions for both chiglitazar doses versus the trial-specific comparator were retained and ranked independently within each trial by their mean percentage contribution across doses. Bootstrap standard errors of contribution percentages were estimated using 2,000 participant resamples within trial-by-treatment strata, refitting the contribution regressions and recalculating the denominator.

#### Molecular associations with IR, adiposity and randomized treatment response

Baseline molecular measurements were expressed in standard-deviation units. Associations with continuous measured log HOMA-IR and BMI were estimated separately, and jointly in mutually adjusted models, with sex, age and trial as covariates. BH correction covered all 148 candidate analytes separately for each exposure and model; displaying the shared 52 analytes did not reduce the correction family. The baseline categorical comparisons regressed each molecular measurement on measured IR status or BMI category, adjusting for sex, age and trial. Normal weight, overweight and obesity were defined as BMI 18.5 to <23, 23 to <25 and ≥25 kg/m^2^, respectively. IR versus non-IR, overweight versus normal weight and obesity versus normal weight contrasts were estimated.

#### Sex-related response heterogeneity

Observed baseline-to-week-24 four-category transitions were tabulated among 387 chiglitazar-treated participants with paired classifications. Subsequent sex analyses focused on the 136 participants with baseline IR+/mol-IR+ status (85 men and 51 women). Conversion to IR−/mol-IR− was analyzed by logistic regression, and mol-HOMA-IR change by OLS with HC3 standard errors. Primary models included sex, baseline measured and molecular HOMA-IR, trial and dose. Additional models included age and baseline log HbA1c, log HDL cholesterol and log triglycerides; further sensitivities added log BMI or PC1–PC5, with complete-case matched comparisons where appropriate. Sex effects on conversion were assessed by likelihood-ratio tests; continuous effects used two-sided *t*-tests. BH correction covered the two endpoints within each covariate set. Adjusted means or probabilities were standardized to the eligible sample’s covariate distribution.

Trial-specific analyses included all treatment arms among participants with baseline double-positive status. Continuous models included sex, treatment arm, their interaction, and baseline measured and molecular HOMA-IR; HC3 F-tests jointly assessed the two interaction terms. Four-category outcomes were modeled with mean-bias-reduced multinomial logistic regression using the same predictors. Joint Wald tests assessed the six sex-by-arm interaction terms, and three-degree-of-freedom Wald tests assessed sex differences within each arm. Bias-reduced binary logistic models of conversion to the double-negative state were also evaluated. BH correction covered four primary interaction tests (two endpoints × two trials), with six within-arm sex tests corrected separately for each endpoint. Binary-conversion interaction tests formed a separate two-test family. As a sensitivity analysis, ordinary multinomial likelihood-ratio interaction tests were calibrated using 1,999 parametric bootstrap simulations under the fitted no-interaction model, holding covariates fixed. The bootstrap *P* value was (b + 1)/(1,999 + 1), where b counted simulated statistics at least as large as the observed statistic; BH adjustment covered the two trial-specific bootstrap tests. Bias-reduced multinomial Wald tests remained the primary inference. The sex analyses were exploratory and were not prespecified in the parent trials.

## Supplementary Figures

**Figure S1.**
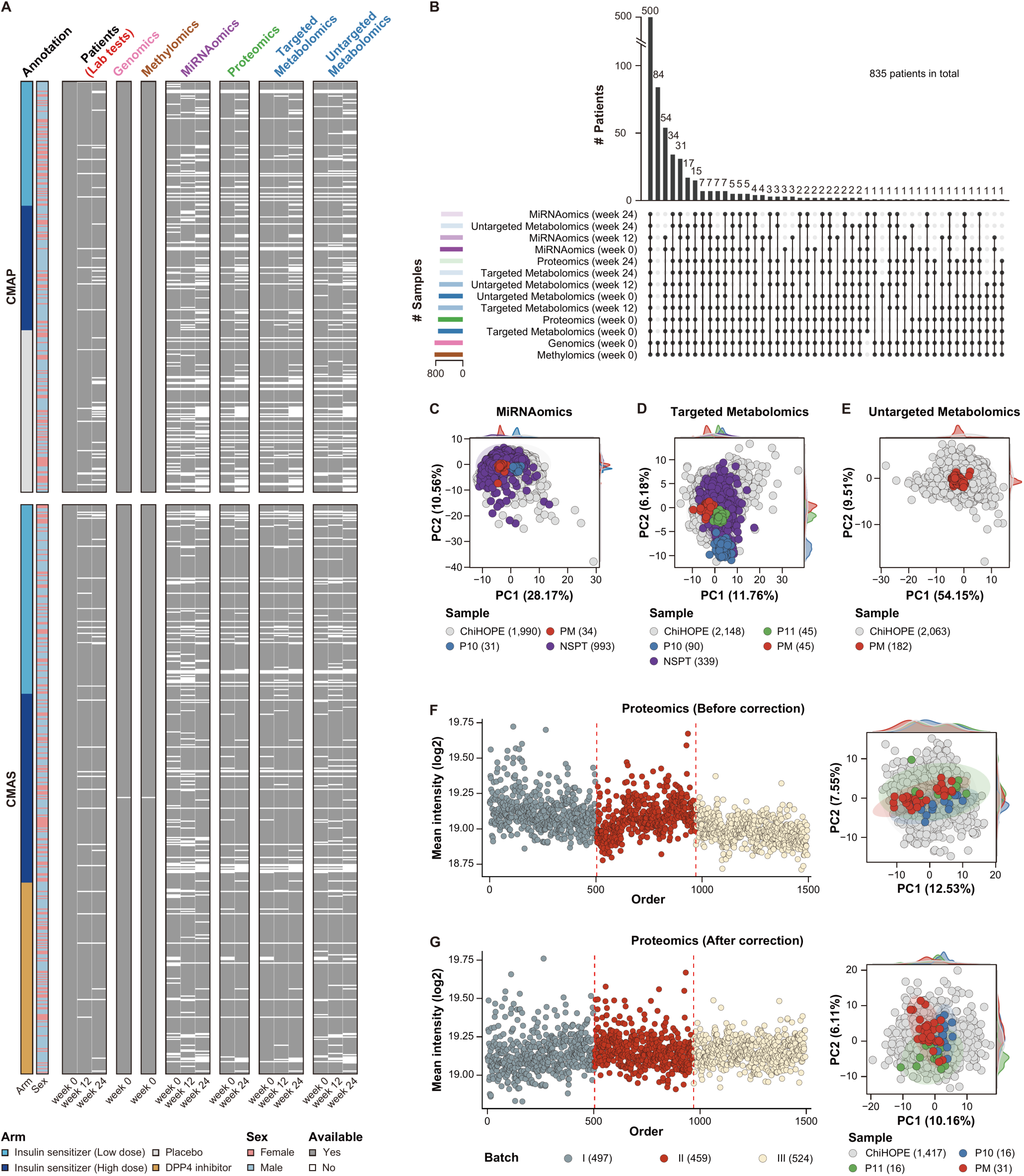
Overview of the longitudinal multi-omics datasets, related to Figure 1. (**A**) Data availability for each patient at different time points. Each row represents a patient (sample), and each column represents a data type at a given time point. (**B**) Number of patients with available data at different time points. (**C**) The 1st (x-axis) and the 2nd (y-axis) principal components (PCs) of the miRNAomic dataset. We collected plasma samples from the ChiHOPE cohort (*n* = 1,990) and the NSPT cohort (*n* = 993). Plasma samples of a healthy individual (P10, *n* = 31) and mixed plasma samples from the ChiHOPE cohort (PM, *n* = 34) were detected for quality control (QC). The variances explained by PC1 and PC2 are labeled in the brackets of the axis titles. (**D**) The PC1 (x-axis) and PC2 (y-axis) of the targeted metabolomic dataset. We collected plasma samples from the ChiHOPE cohort (*n* = 2,148) and the NSPT cohort (*n* = 339). Plasma samples of two healthy individuals (P10, *n* = 90; P11, *n* = 45) and the PM samples (*n* = 45) were detected for QC. (**E**) The PC1 (x-axis) and PC2 (y-axis) of the untargeted metabolomic dataset. Plasma samples were collected from the ChiHOPE cohort (*n* = 2,063). The PM samples (*n* = 182) were detected for QC. (**F**-**G**) The proteomic datasets before (**F**) and after (**G**) batch-effect correction. Scatter plots (left) show the log-transformed mean intensity (y-axis) of samples with different loading orders (x-axis), which can be grouped into three batches (I: *n* = 497; II: *n* = 459; and III: *n* = 524). The scatter plots (right) show PC1 (x-axis) and PC2 (y-axis). Plasma samples were collected from the ChiHOPE cohort (*n* = 1,417). The P10 (*n* = 16), P11 (*n* = 16) and PM (*n* = 31) samples were detected for QC.

**Figure S2.**
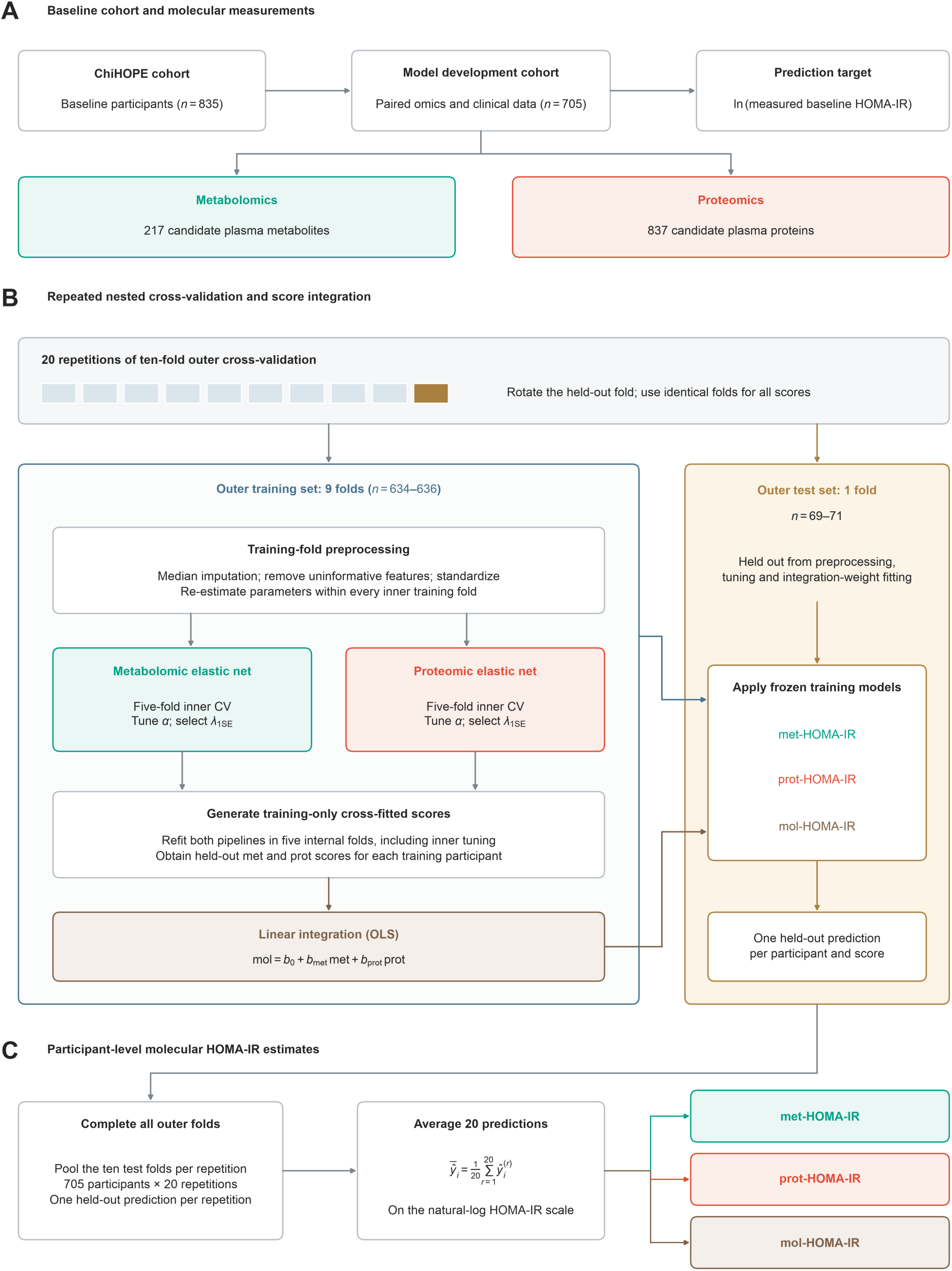
Development of molecular HOMA-IR estimates using repeated nested cross-validation. (**A**) Baseline molecular measurements included 217 candidate metabolites and 837 candidate proteins in 705 ChiHOPE participants with paired metabolomic, proteomic and clinical measurements. The prediction target was natural-log-transformed measured HOMA-IR. (**B**) Models were developed using 20 repetitions of ten-fold outer cross-validation, with identical participant splits across all three molecular estimates. Each outer training set comprised 634–636 participants, and the remaining 69–71 participants formed the held-out test set. Targeted metabolomic inputs had already undergone platform-level half-minimum imputation across all measured samples and logarithmic transformation before model development; this upstream processing was not repeated within cross-validation folds. Within each training split, any remaining missing molecular values were imputed using training-set medians, features with insufficient observations or negligible variability were removed, and retained features were standardized using training-set parameters. We re-estimated all model-stage preprocessing parameters within inner training folds. Separate elastic-net regression models were fitted for metabolomics and proteomics to derive met-HOMA-IR and prot-HOMA-IR, respectively. Hyperparameters were selected using five-fold inner cross-validation: α was selected by minimizing the inner cross-validation mean squared error, and λ was selected using the one-standard-error rule. To derive mol-HOMA-IR, we performed an additional five-fold cross-fitting procedure within each outer training set to generate metabolomic and proteomic predictions, repeating model-stage preprocessing and inner tuning within each internal training subset. We combined these cross-fitted predictions using OLS linear regression to predict measured log HOMA-IR. The fitted single-omics models and integration weights were then applied to the outer test set. Outer test participants were excluded from model-stage preprocessing estimation, hyperparameter tuning and integration-weight estimation. Molecular prediction models did not include clinical covariates or adiposity residualization. (**C**) Predictions from the ten outer test folds were pooled within each repetition, yielding one held-out prediction per participant for each molecular estimate. We averaged each participant’s 20 held-out predictions on the natural-log HOMA-IR scale to obtain the final met-HOMA-IR, prot-HOMA-IR, and mol-HOMA-IR estimates. *n*, number of participants; CV, cross-validation; OLS, ordinary least squares; SE, standard error.

**Figure S3.**
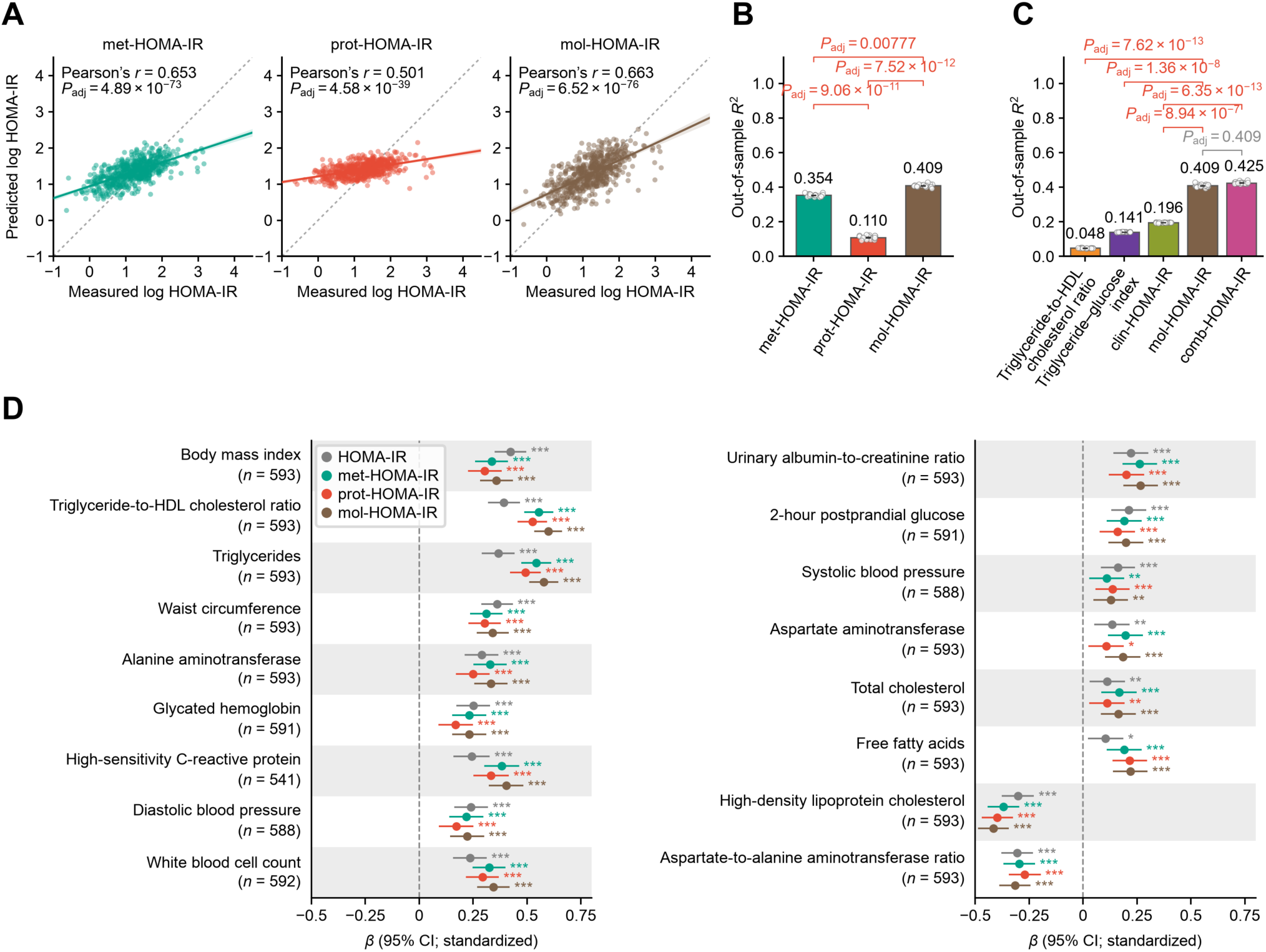
Molecular HOMA-IR performance and clinical associations at week 24. (**A**) Correlation between the measured and predicted log-transformed HOMA-IR in 593 participants at W24. The solid line is the OLS linear regression line with a 95% confidence interval, and the dotted line is measured log-transformed HOMA-IR = predicted log-transformed HOMA-IR. *P*_adj_: BH-adjusted *P value* of a two-sided Pearson’s correlation test across the three categories. (**B-C**) Model performance of each fitted HOMA-IR model. For each repetition, out-of-sample *R*^2^ was calculated by pooling held-out predictions across all ten outer folds. Bars show mean ± standard error (SE) across 20 repetitions. Numbers above bars indicate mean *R*^2^. *P*_adj_: BH-adjusted *P value* of a two-sided Nadeau–Bengio corrected paired t-test on MSE differences across the 200 outer test folds. BH correction covered three pairwise comparisons in B and all ten pairwise comparisons in C; five selected comparisons are displayed in C. (**D**) Association between molecular HOMA-IR estimates and clinical measures. For each of the 18 clinical measures, *a β*-coefficient was estimated using an OLS linear regression model with each measure as the dependent variable and the measured or molecular HOMA-IR estimate as the predictor. Models were adjusted for sex, age and clinical trial. Associations used available observations from the 593-participant week-24 cohort. Multiple testing correction was performed using the BH method across 72 clinical regressions (18 measures × four HOMA-IR types). *n*, number of assessed participants. Data: estimates with 95% confidence intervals. *adjusted *P* < 0.05, **adjusted *P* < 0.01, ***adjusted *P* < 0.001. All exact values of test summaries are found in Source Data.

**Figure S4.**
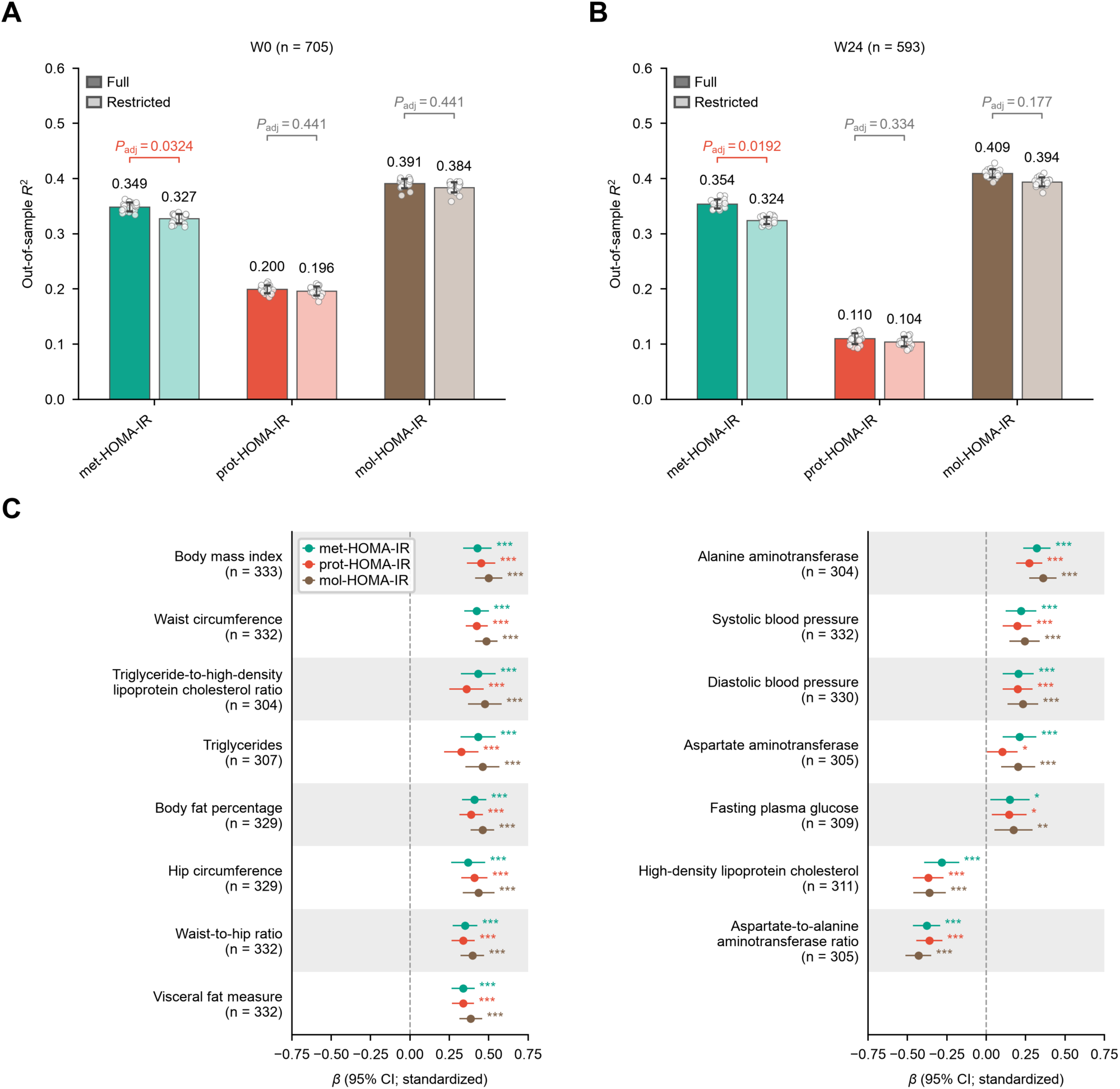
Performance and external clinical associations of restricted molecular HOMA-IR models. (**A-B**) Model performance of each fitted full and restricted HOMA-IR model at baseline (**A**) and W24 (**B**). Restricted models used 202 metabolites and 754 proteins shared with NSPT. For each repetition, out-of-sample *R*^2^ was calculated by pooling held-out predictions across all ten outer folds. Bars show mean R^2^ across 20 repetitions; error bars indicate the SD across repetitions. Numbers above bars indicate mean *R*^2^. *P*_adj_: BH-adjusted *P value* of a*n* approximate two-sided Nadeau–Bengio corrected paired t-test on MSE differences across the 200 outer test folds, with BH correction across the three full-versus-restricted comparisons separately at each visit. (**C**) Associations between restricted molecular HOMA-IR estimates and 15 clinical measures in NSPT (*n* = 333). Final models were fitted using all 705 ChiHOPE baseline participants and frozen before application to NSPT. For each of the 15 clinical measures, *a β*-coefficient was estimated using an OLS linear regression model with each measure as the dependent variable and the molecular HOMA-IR estimate as the predictor adjusted for sex and age. We used HC3 heteroscedasticity-robust standard errors and two-sided t-tests for inference. Multiple testing correction was performed using the BH method across 45 clinical regressions (15 measures × three HOMA-IR types). *n*, number of assessed participants. Data: estimate with 95% confidence interval. *adjusted *P* < 0.05, **adjusted *P* < 0.01, ***adjusted *P* < 0.001. All exact values of test summaries are found in Source Data.

**Figure S5.**
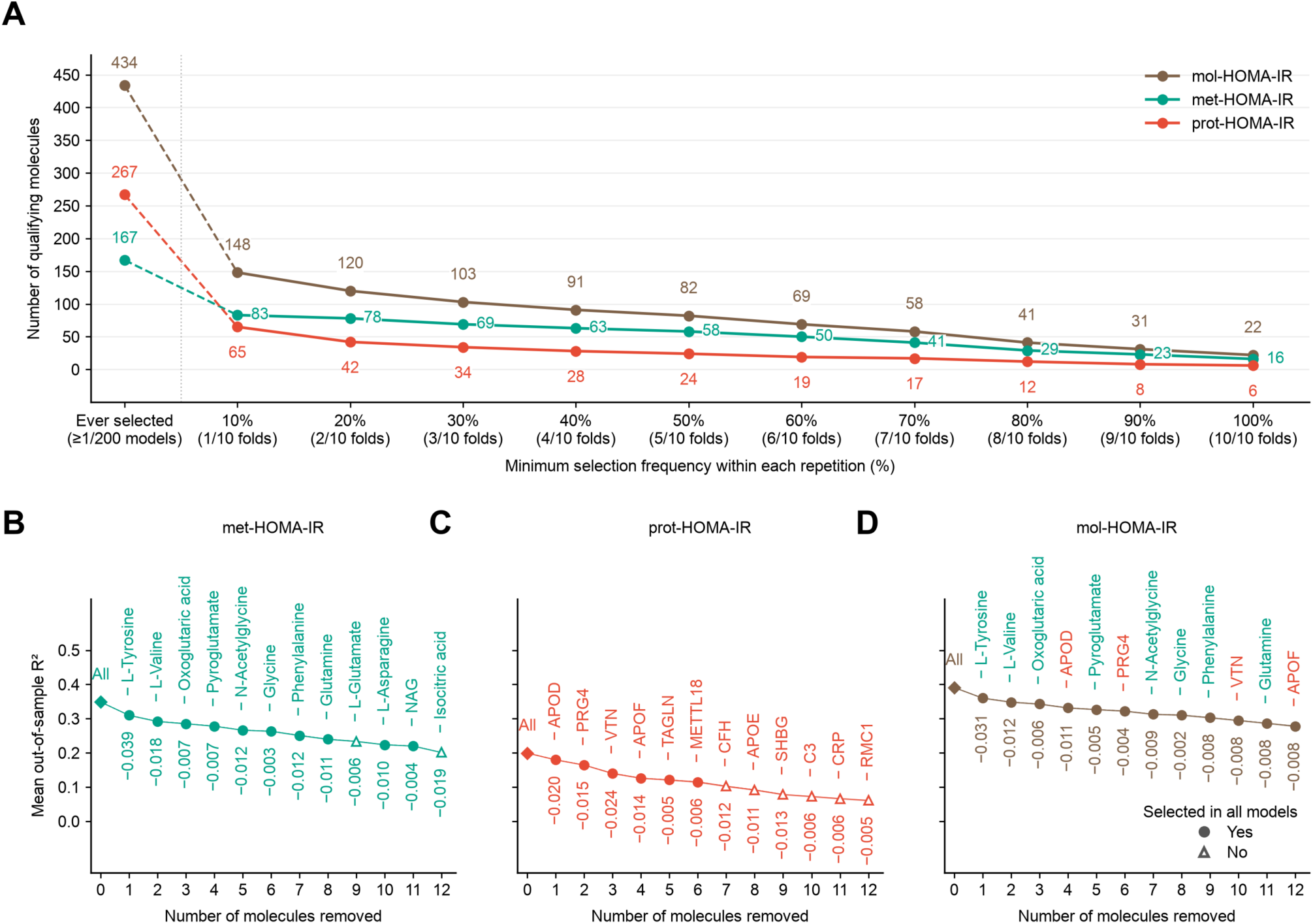
Feature-selection stability and sensitivity to sequential feature removal. (**A**) Numbers of features meeting selection-frequency thresholds within every repetition. “Ever selected” denotes selection in at least one of 10 folds in one of 20 repetitions; percentage thresholds require selection in the specified number of folds in each of the 20 repetitions. (**B**-**D**) Mean out-of-sample *R*^2^ during sequential feature removal from met-HOMA-IR (**B**), prot-HOMA-IR (**C**) and mol-HOMA-IR (**D**). At each iteration, the eligible feature with the largest absolute mean coefficient across the outer models was removed. Models were retuned and refitted after each deletion. “All” denotes the undeleted model; labels identify the removed analyte, and numbers below the curves indicate the *R*^2^ change from the preceding iteration.

**Figure S6.**
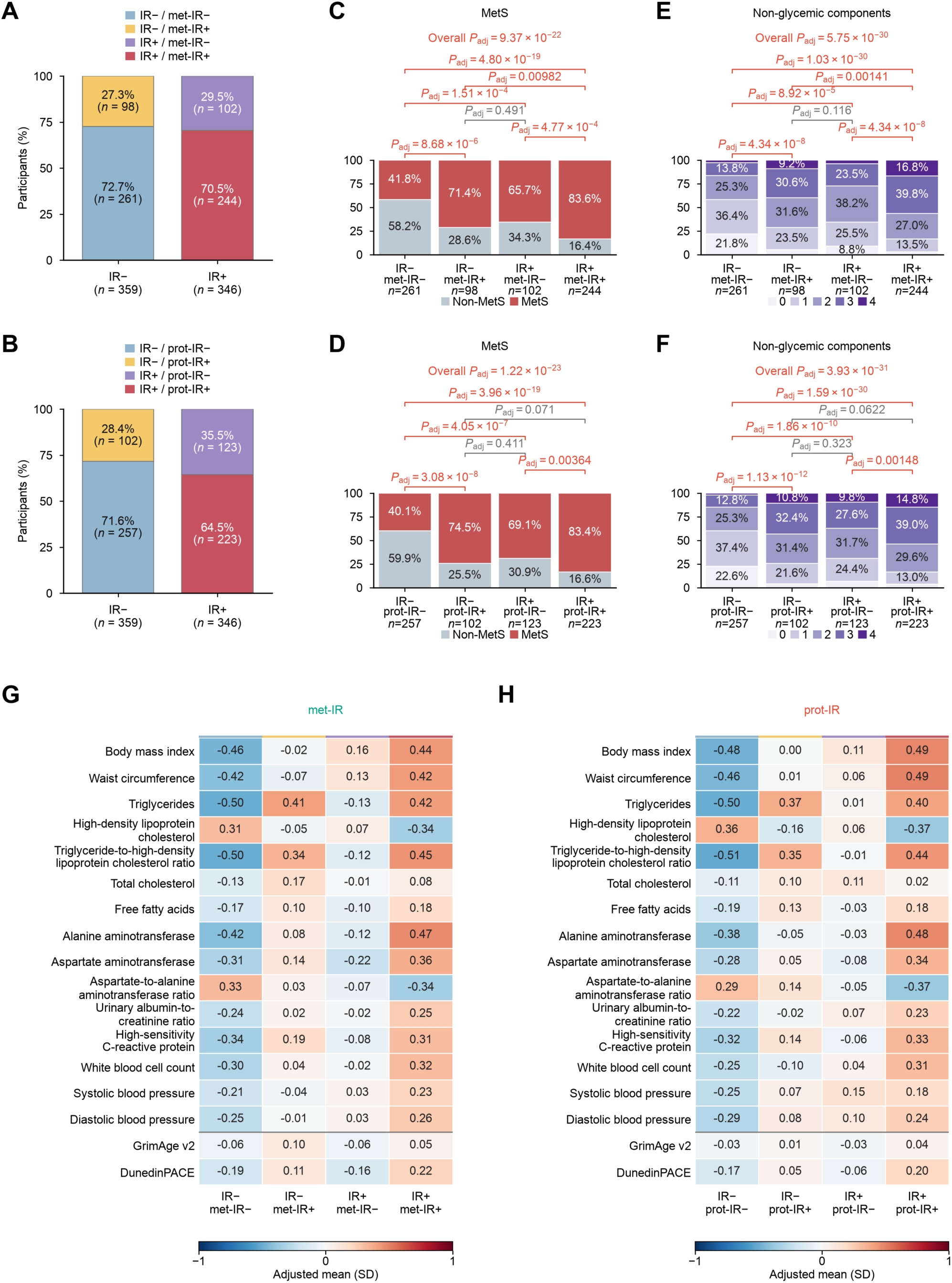
Met-IR and prot-IR captured metabolic heterogeneity in classical IR classes. Analyses used available observations from the 705-participant baseline cohort. (**A-B**) Distribution of met-IR (**A**) and prot-IR (**B**) status within measured IR− and IR+ groups. Measured and molecular IR were defined using sex-specific HOMA-IR thresholds of ≥4.31 in men and ≥4.51 in women. (**C-F**) Observed distributions of metabolic syndrome (MetS; C**-**D) and the number of non-glycemic MetS components (**E-F**), using met-IR classifications in C and E and prot-IR classifications in D and F. The four non-glycemic components were abdominal obesity, elevated blood pressure, elevated triglycerides and low HDL cholesterol. (**G-H**) Covariate-adjusted means of 15 clinical measures and two epigenetic aging measures across the four groups defined using met-IR (**G**) or prot-IR (**H**), expressed in standard-deviation units. We log-transformed clinical measures before standardization. Four-group models adjusted for sex, age and clinical trial, with additional blood-cell adjustment for epigenetic aging measures. Adjusted means were standardized to the covariate distribution of the eligible analysis sample. Overall and pairwise differences were assessed using OLS F- and t-tests for continuous measures, logistic likelihood-ratio and Wald tests for MetS, and OLS with HC3 robust F- and t-tests for component counts. For continuous clinical measures, BH adjustment covered 54 overall tests and 324 pairwise comparisons across three molecular classifications. For MetS-related outcomes, adjustment covered 18 overall tests and 108 pairwise comparisons across six outcomes and three classifications.

**Figure S7.**
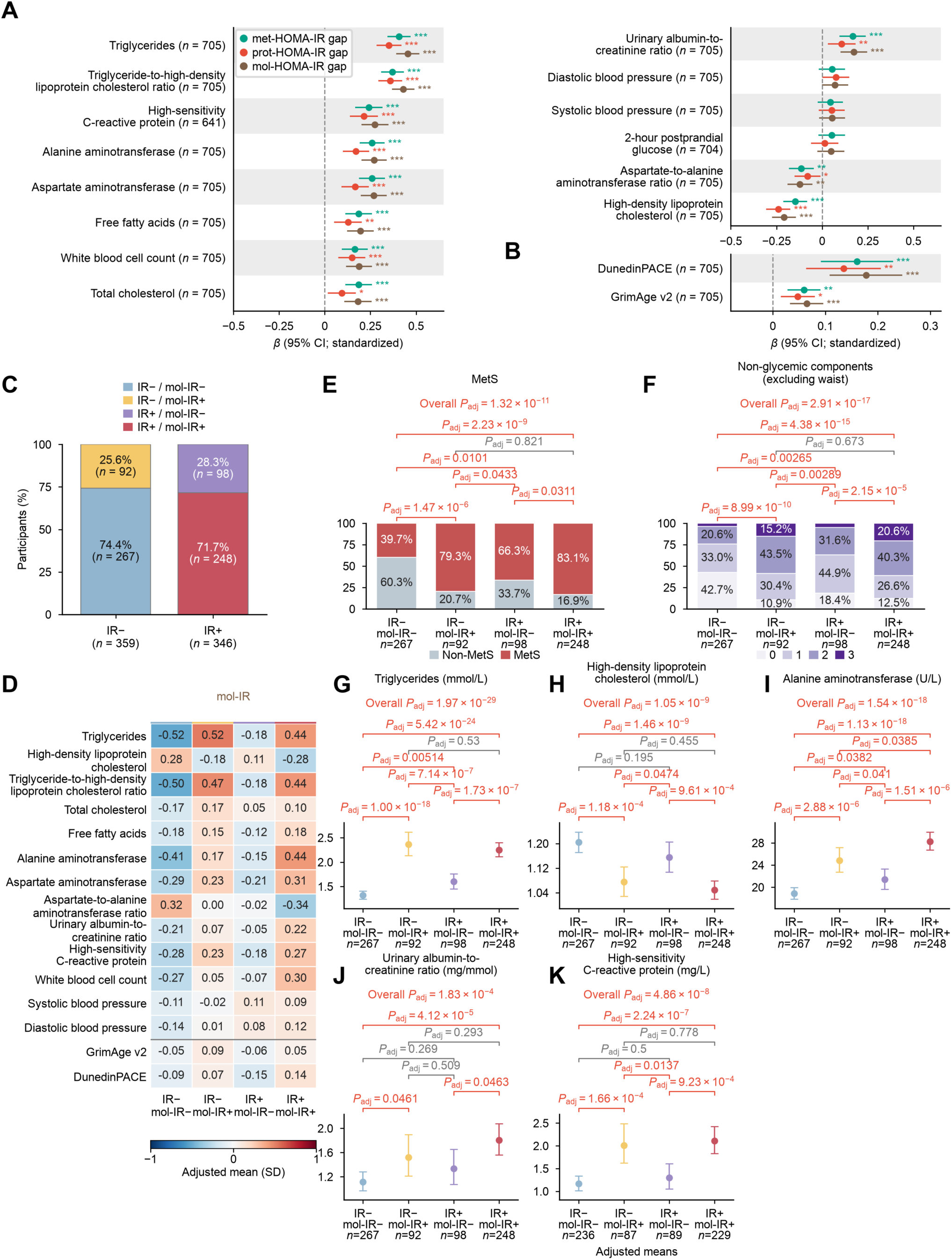
Baseline molecular IR associations and four-group differences after adjustment for BMI and waist circumference. Analyses used available observations from the 705-participant baseline cohort. (**A-B**) Baseline associations between molecular HOMA-IR gaps and clinical measures (**A**) and epigenetic aging measures (**B**). Gaps were residual deviations of predicted log HOMA-IR from its expected value given measured log HOMA-IR, calibrated in outer training sets and averaged across 20 cross-validation repetitions. OLS models regressed each of 16 clinical and 11 epigenetic aging measures on each gap. Models were adjusted for BMI, waist circumference, log-transformed HOMA-IR, sex, age and clinical trial, and models for epigenetic aging measures additionally adjusted for estimated blood-cell composition. Multiple testing correction was performed using the BH method across 48 clinical regressions (16 measures × three gaps) and 33 epigenetic aging regressions (11 measures × three gaps). *n*, number of assessed participants. Data: estimate with 95% confidence interval. *adjusted *P* < 0.05, **adjusted *P* < 0.01, ***adjusted *P* < 0.001. All exact values of test summaries are found in Source Data. (**C**) Distribution of mol-IR status within measured IR− and IR+ groups. Measured and molecular IR were defined using sex-specific HOMA-IR thresholds of ≥4.31 in men and ≥4.51 in women. (**D**) Covariate-adjusted means of 13 clinical measures and two epigenetic aging measures across the four groups, expressed in standard-deviation units. We log-transformed clinical measures before standardization. (**E-F**) Observed distributions of metabolic syndrome (MetS; **E**) and the number of non-glycemic MetS components (**F**). MetS retained its original definition, including abdominal obesity. The non-glycemic component count in F excluded waist circumference and counted elevated blood pressure, elevated triglycerides and low HDL cholesterol (range, 0–3). (**G-K**) Adjusted geometric means and 95% confidence intervals for triglycerides (**G**), HDL cholesterol (**H**), alanine aminotransferase (**I**), urinary albumin-to-creatinine ratio (**J**) and high-sensitivity C-reactive protein (**K**). Four-group models adjusted for BMI, waist circumference, sex, age and clinical trial, with additional blood-cell adjustment for epigenetic aging measures. Adjusted means were standardized to the covariate distribution of the eligible analysis sample. Overall and pairwise differences were assessed using OLS F- and t-tests for continuous measures, logistic likelihood-ratio and Wald tests for MetS, and OLS with HC3 robust F- and t-tests for component counts. BMI and waist circumference were omitted as clinical outcomes. For the remaining 16 clinical measures, BH adjustment covered 48 overall tests and 288 pairwise comparisons across three molecular classifications. For epigenetic aging measures, adjustment covered 33 overall tests and 198 pairwise comparisons across 11 measures and three classifications. For MetS-related outcomes, adjustment covered 15 overall tests and 90 pairwise comparisons across five outcomes (MetS, the three-component count, elevated blood pressure, elevated triglycerides and low HDL cholesterol) and three classifications; abdominal obesity was omitted as a separate endpoint.

**Figure S8.**
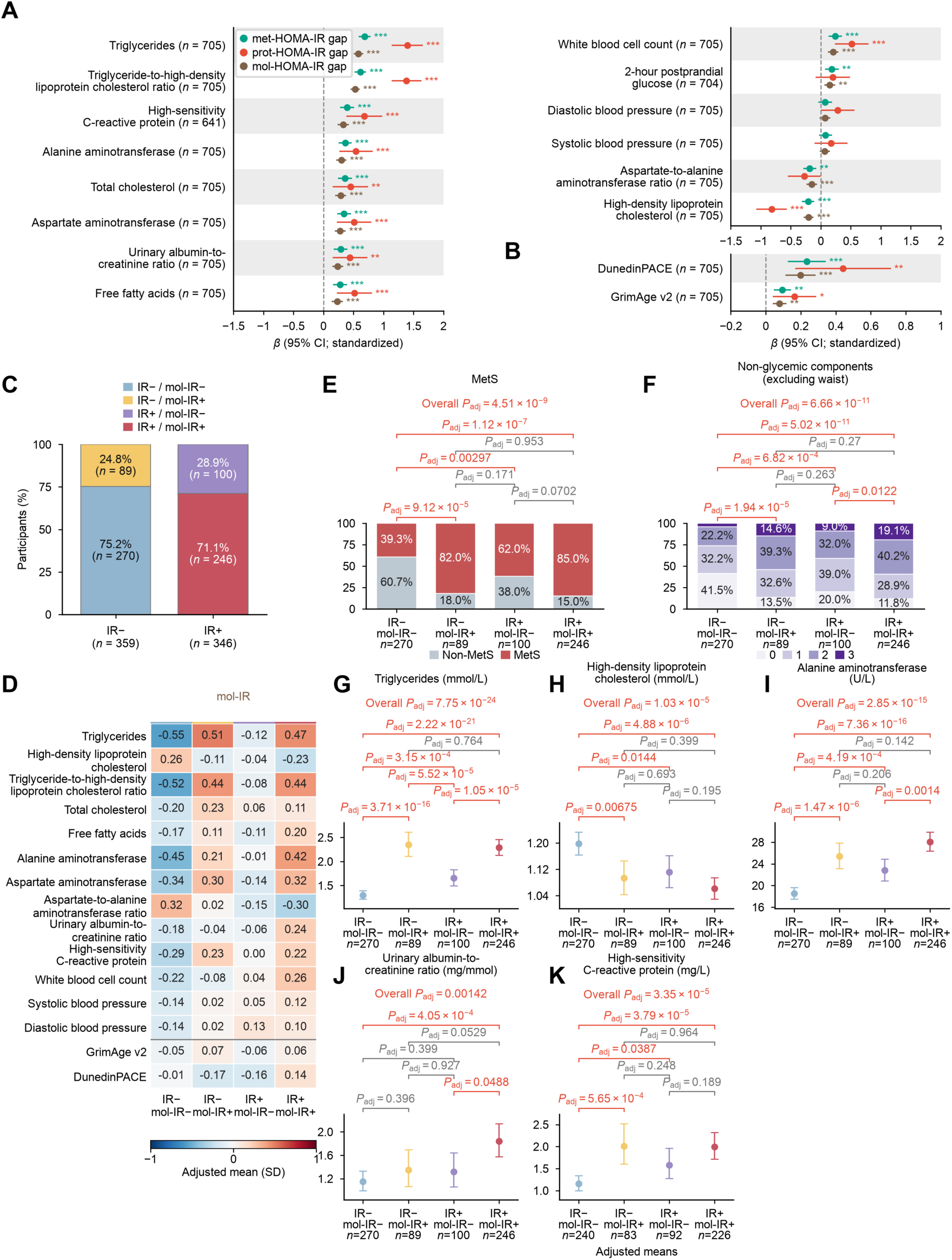
Baseline molecular IR heterogeneity after adjustment for BMI and waist circumference, using models trained with BMI as an unpenalized covariate. Molecular HOMA-IR estimates were derived from models retrained with BMI as an unpenalized covariate in 705 baseline participants. All panels show baseline associations or distributions. (**A**–B) Associations between molecular HOMA-IR gaps and clinical measures (A) and epigenetic aging measures (B). Gaps were calculated using calibration models fitted within the outer training sets and were averaged across 20 cross-validation repetitions. For each of 16 clinical measures, excluding BMI and waist circumference as outcomes, we fitted an OLS linear regression model with the standardized log-transformed clinical measure as the dependent variable and the standardized molecular gap as the predictor, adjusting for BMI, waist circumference, measured log HOMA-IR, sex, age and clinical trial. Epigenetic aging models analyzed 11 outcomes on their original scales and additionally adjusted for estimated blood-cell composition; coefficients were divided by the outcome SD for display. BH adjustment covered 48 clinical regressions (16 measures × three gaps) and 33 epigenetic aging regressions (11 measures × three gaps), separately. Fourteen clinical measures and two epigenetic aging measures are displayed. Data: standardized *β*-coefficient with 95% confidence interval. (**C**) Distribution of mol-IR status within measured IR− and IR+ groups. Measured and molecular IR were defined using sex-specific HOMA-IR thresholds of ≥4.31 in men and ≥4.51 in women, applied as log thresholds to molecular scores. (**D**) Covariate-adjusted means of 13 clinical measures and two epigenetic aging measures across the four groups, expressed in standard-deviation units. We log-transformed clinical measures before standardization. (**E**–F) Observed distributions of metabolic syndrome (MetS; E) and the number of non-glycemic components excluding waist circumference (**F**). MetS retained its original definition, including abdominal obesity. The component count included elevated blood pressure, elevated triglycerides and low HDL cholesterol (range, 0–3). (**G–K**) Adjusted geometric means and 95% confidence intervals for triglycerides (**G**), HDL cholesterol (**H**), alanine aminotransferase (**I**), urinary albumin-to-creatinine ratio (**J**) and high-sensitivity C-reactive protein (**K**). Four-group models adjusted for BMI, waist circumference, sex, age and clinical trial, with additional blood-cell adjustment for epigenetic aging measures. This downstream adjustment was in addition to including BMI when training the molecular models. Adjusted means were standardized to the covariate distribution of the eligible analysis sample. Overall and pairwise differences were assessed using OLS F- and two-sided t-tests for continuous measures, logistic likelihood-ratio and Wald tests for MetS, and OLS with HC3 robust F- and t-tests for component counts. For continuous clinical measures, BH adjustment covered 48 overall tests and 288 pairwise comparisons across 16 measures and three molecular classifications. For epigenetic aging measures, BH adjustment covered 33 overall tests and 198 pairwise comparisons across 11 measures and three molecular classifications. For MetS-related outcomes, adjustment covered 15 overall tests and 90 pairwise comparisons across five outcomes (MetS, the three-component count, elevated blood pressure, elevated triglycerides and low HDL cholesterol) and three classifications; abdominal obesity was omitted as a separate endpoint. *n*, number of assessed participants, varying by outcome availability as indicated in the panels. *Adjusted *P* < 0.05, **adjusted *P* < 0.01, ***adjusted *P* < 0.001. All exact values of test summaries are found in Source Data.

**Figure S9.**
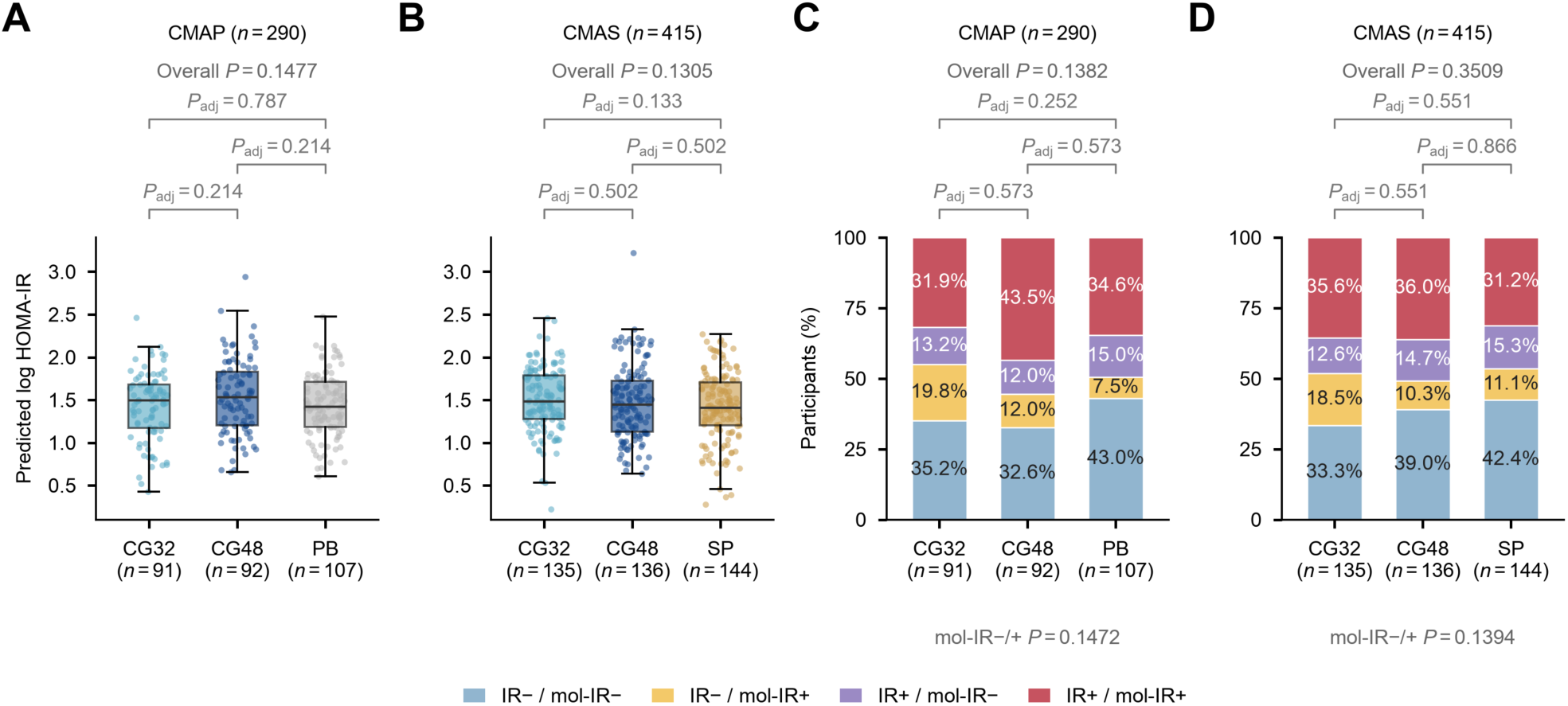
Baseline molecular HOMA-IR and mol-IR classifications by treatment group. (**A**-**B**) Baseline mol-HOMA-IR estimates in CMAP (A; n=290) and CMAS (B; n=415). Points represent participants; boxes indicate medians and interquartile ranges, and whiskers extend to the most extreme observations within 1.5 interquartile ranges of the box. All participant points are shown. Scores are the mean held-out natural-log predictions across 20 cross-validation repetitions. Overall P values are from Welch one-way ANOVA. (**C**-**D**) Observed proportions in the four categories defined by measured IR and mol-IR in CMAP (C) and CMAS (D). Sex-specific HOMA-IR thresholds of 4.31 for men and 4.51 for women were applied to both measured HOMA-IR and molecular estimates (as log thresholds for log scores). Four-class P values compare the full four-category distribution; mol-IR−/+ P values compare marginal molecular IR status. Both use Pearson chi-square tests. All tests compare the three treatment groups within each trial. Overall P values are from unadjusted exploratory omnibus tests. Brackets indicate pairwise comparisons, using Welch t-tests in A, B and Pearson chi-square tests of the complete four-class distribution in C, D. Pairwise P values (P_adj) are Holm-adjusted over the three arm pairs separately within each trial and outcome; no covariate adjustment was applied. Binary overall P values below C, D are supplementary and do not correspond to the four-class brackets. n denotes the number of participants per treatment arm. Lack of statistical significance does not demonstrate equivalence.

**Figure S10.**
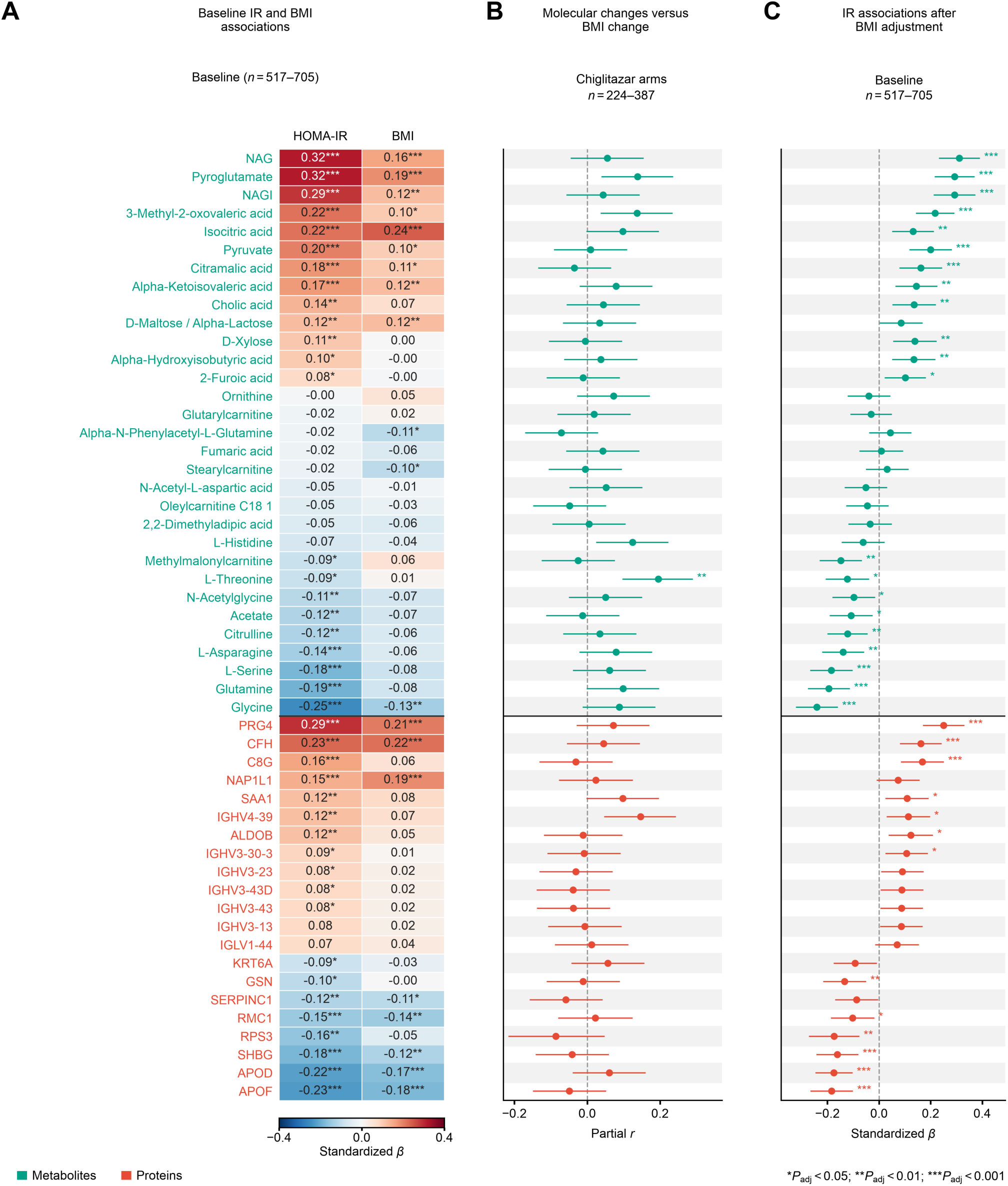
Associations of shared treatment-associated molecular contributors with insulin resistance and adiposity. Analyses included all 52 analytes (31 metabolites and 21 proteins) contributing to lower mol-HOMA-IR with both chiglitazar doses relative to placebo in CMAP and relative to sitagliptin in CMAS. Selection required negative contributions in all four dose–control comparisons, without an association-significance filter. These analytes were drawn from the 148 candidates selected in at least one of ten outer-fold models in every cross-validation repetition. (**A**) Baseline associations of molecular levels with measured log-transformed HOMA-IR and BMI in the 705-participant baseline cohort. Separate OLS models regressed each standardized molecular level on standardized log HOMA-IR or standardized BMI, adjusting for sex, age, clinical trial. Heatmap cells show standardized *β*-coefficients; 41 analytes were associated with measured HOMA-IR, 20 with BMI and 18 with both. (**B**) Partial correlations between molecular changes and BMI changes in the chiglitazar arms pooled across both trials, adjusting for baseline molecular level, baseline BMI, sex, age and trial-by-dose stratum. Data: partial correlation estimate with approximate 95% confidence interval. (**C**) Baseline associations with measured log-transformed HOMA-IR after additional adjustment for BMI, using OLS models with standardized molecular level as the dependent variable, standardized log HOMA-IR as the predictor and the baseline covariates in A. Data: standardized *β*-coefficient with 95% confidence interval. Available-case sample sizes varied by analyte: *n* = 517–705 for A and C and *n* = 224–387 for B. Molecular levels were standardized using available observations in the eligible baseline sample; the same molecular transformation was used at week 24. Rows are aligned across A–C and ordered by baseline HOMA-IR coefficient within omics category. For A–C, two-sided t-test *P* values were BH-adjusted across the original 148 candidate analytes separately within exposure, model variant and analysis subgroup; adjustment was not recalculated for the 52 displayed analytes. *Adjusted *P*<0.05, **adjusted *P*<0.01, ***adjusted *P*<0.001. Confidence intervals are not multiplicity-adjusted. *n*, number of assessed participants. All exact values, analyte-specific sample sizes and test summaries are provided in the accompanying source-data tables.

**Figure S11.**
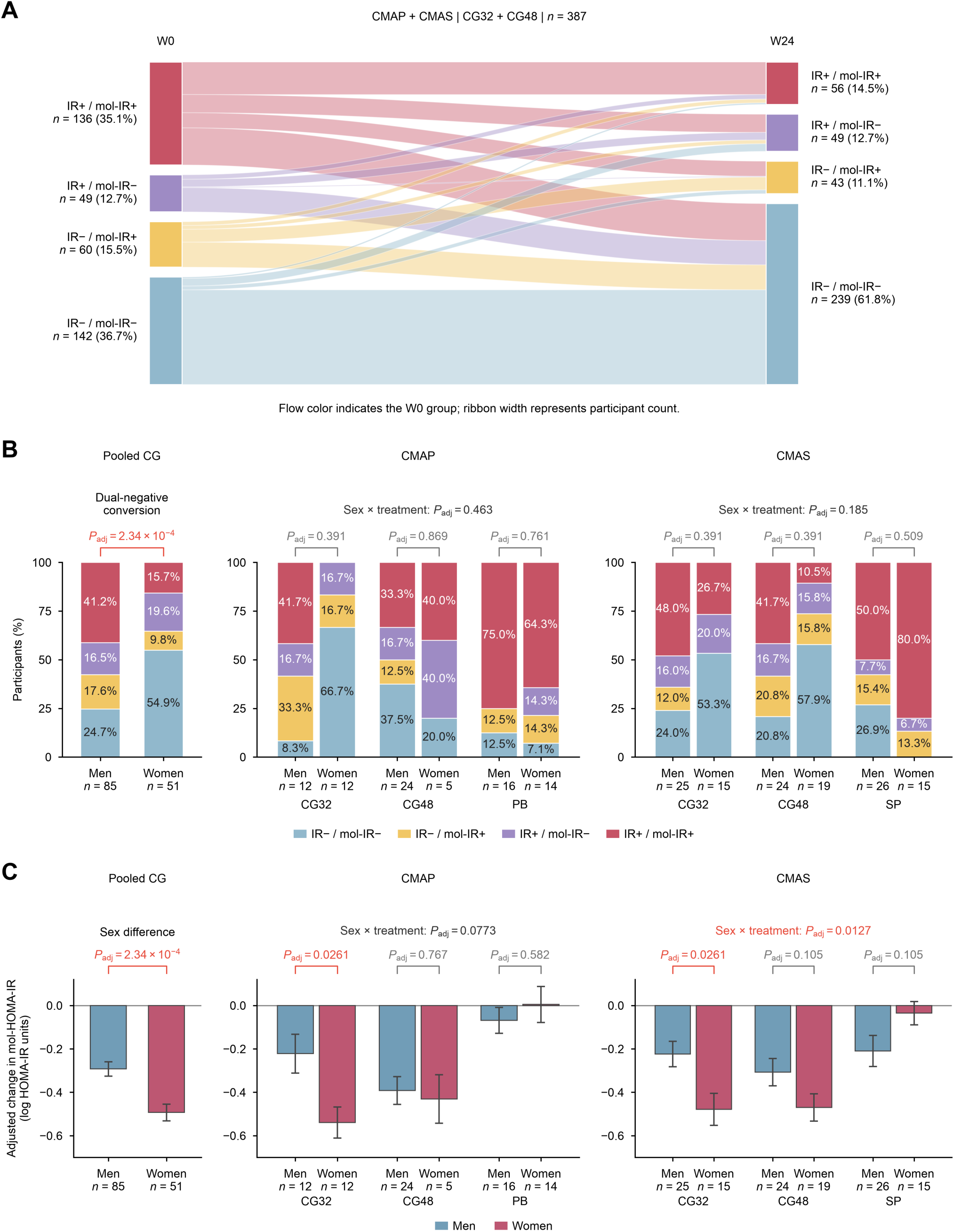
Molecular IR responses by sex among participants with baseline IR+/mol-IR+ status. (**A**) Transitions from baseline (W0) to week-24 (W24) measured IR/mol-IR classifications among 387 chiglitazar-treated participants from the 705-participant modeling cohort with paired classifications. Node heights and ribbon widths are proportional to participant counts; ribbon colors indicate the baseline category. Numbers and percentages next to nodes describe the proportion of all 387 participants at each visit. (**B**) Observed week-24 four-category proportions by sex, shown for pooled chiglitazar treatment (left; *n* = 136), the three treatment arms in CMAP (middle; *n* = 83), and the three treatment arms in CMAS (right; *n* = 124). All participants in B and C were IR+/mol-IR+ at baseline. The pooled CG sample includes 85 men and 51 women; trial-specific plots additionally include the corresponding placebo or sitagliptin control. Percentages in B are calculated within each bar. (**C**) Covariate-adjusted changes in mol-HOMA-IR estimates in the same three analysis populations. Data: adjusted mean ± standard error (SE), using HC3 covariance. Changes are week 24 minus baseline in log-HOMA-IR units; negative values indicate a decrease. Pooled CG models include sex, baseline measured log HOMA-IR, baseline mol-HOMA-IR, trial and dose. The annotation in the pooled part of B tests conversion to IR−/mol-IR− versus all other outcomes using binary logistic regression and a likelihood-ratio test; the pooled part of C uses OLS with an HC3 t-test. BH adjustment covers these two pooled sex-comparison endpoints. Trial-specific models include sex, three-level treatment, their interaction, and baseline measured log HOMA-IR and mol-HOMA-IR. Adjusted means are standardized to the covariate distribution of the corresponding eligible analysis sample. Trial-specific four-category outcomes in B use mean-bias-reduced multinomial logistic models and joint Wald tests (6 df for sex-by-treatment interaction; 3 df for a within-arm sex contrast); these tests differ from the binary-conversion test shown in the pooled CG part of B. Continuous outcomes use OLS HC3 F-tests (2 df for interaction) and t-tests (within-arm sex contrasts). BH adjustment covers four primary interaction tests (two outcomes × two trials), and separately six within-arm sex contrasts per outcome. The categorical interaction inference in CMAS was method-sensitive: the primary bias-reduced Wald test was not significant (unadjusted *P* = 0.139), whereas the 1,999-draw parametric-bootstrap likelihood-ratio sensitivity test gave *P* = 0.0020. These sensitivity results and binary-conversion arm-specific tests are provided in the accompanying tables. Small segments in B are not labeled when text would overlap. *n*, number of participants. CG32 and CG48, chiglitazar 32 and 48 mg; PB, placebo; SP, sitagliptin. Exact values and complete model specifications are provided in the accompanying data files.

## Notes

### Clinical Trial

CMAP: NCT02121717; CMAS: NCT02173457.

### Author Declarations

The CMAP trial (NCT02121717) was approved by the Medical Ethics Committee of Peking University People's Hospital, and the CMAS trial (NCT02173457) was approved by the Ethics Committee of Shanghai Sixth People's Hospital. Ethics approval was also obtained from the institutional review boards of the participating study centers. Written informed consent covered blood collection and subsequent multi-omics research. The NSPT cohort was approved by the Ethics Committee of Human Genetic Resources of the School of Life Sciences, Fudan University, Shanghai, China (approval number 14117).

